# Predicting patient-reported outcomes following lumbar spine surgery: development and external validation of multivariable prediction models

**DOI:** 10.1101/2022.02.15.22270980

**Authors:** Monika Halicka, Martin Wilby, Rui Duarte, Christopher Brown

## Abstract

**Background:** This study aimed to develop and externally validate prediction models of spinal surgery outcomes based on a retrospective review of a prospective clinical database, uniquely comparing multivariate regression and machine learning approaches, and identifying the most important predictors.

**Methods:** Outcomes were change in back and leg pain intensity and Core Outcome Measures Index (COMI) from baseline to the last available postoperative follow-up (3-24 months), defined as minimal clinically important change (MCID) and continuous change score. Eligible patients underwent lumbar spine surgery for degenerative pathology between 2011 and 2021. Data were split by surgery date into development (N=2691) and validation (N=1616) sets. Multivariate logistic and linear regression, and random forest classification and regression models, were fit to the development data and validated on the external data.

**Results:** All models demonstrated good calibration in the validation data. Discrimination ability (area under the curve) for MCID ranged from 0.63 (COMI) to 0.72 (back pain) in regression, and from 0.62 (COMI) to 0.68 (back pain) in random forests. The explained variation in continuous change scores spanned 16%-28% in linear, and 15%-25% in random forests regression. The most important predictors included age, baseline scores on the respective outcome measures, type of degenerative pathology, previous spinal surgeries, smoking status, morbidity, and duration of hospital stay.

**Conclusions:** The developed models appear robust and generalisable across different outcomes and modelling approaches but produced only borderline acceptable discrimination ability, suggesting the need to assess further prognostic factors. External validation showed no advantage of the machine learning approach.

## 1. Background

Chronic back pain is the single greatest cause of years lived with disability worldwide [1] with annual direct healthcare costs in the UK of £1,632 million [2]. Spinal surgery is the largest component of healthcare expenditure for managing low back pain (costing £5000-£10,000 per patient) and its rate has doubled over 15-years from 2.5 to 4.9 per 10,000 adults [3]. However, success rates of lumbar spine surgery are highly variable - only about 60% of patients achieve reductions in pain of at least a minimal clinically important difference (MCID), and 1/5 experience persistent long-term pain after surgery [3–5].

Reliable predictive factors could maximise patient benefit and cost-effectiveness of surgery. Although several systematic reviews concluded that medical, sociodemographic, and psychological factors are linked to improvement in pain and disability following lumbar spine surgery [6–10], we lack clear clinical guidelines on the reliable predictors [11] and predictive factors are rarely formally documented.

Statistical prediction models enable probabilistic estimation of treatment outcome given a set of preoperative patient data. Previously developed regression-based models demonstrated good ability to discriminate between patients who did and did not achieve MCID in pain or disability after lumbar spine surgery [12–16]. However, performance of the existing prediction models has been rarely quantified in patient data other than that used to develop them. In the few available examples, external validation revealed poorer discrimination ability in the new data [17,18], or poor calibration leading to under- and overestimation of outcome probabilities [19].

Machine learning has been found to improve predictive performance relative to regression [20], including for spinal surgery outcomes [21–23]. This may stem from the ability to capture nonlinear or interactive effects, often characteristic of clinical data. Conversely, machine learning algorithms are prone to overfitting smaller datasets and comparisons to standard regression have lacked external validation; thus, such predictions may not extrapolate well to new data. External validation is rare but necessary before any prediction models can be implemented in clinical practice [24,25].

Our primary objective was to develop and externally validate prediction models of patient-reported spinal surgery outcomes based on routinely collected prospective data, for the first time comparing the performance of multivariate regression and machine learning approaches, the latter hypothesised to improve prediction accuracy. Secondarily, we identify the most relevant predictors from the available medical and patient-reported information, since existing models (particularly those from the US [14–16,21]) may not translate well to UK cohorts due to different healthcare systems, type of data recorded, and cultural and demographic differences.

## 2. Methods

The present article follows the Transparent Reporting of a multivariable prediction model for Individual Prognosis Or Diagnosis (TRIPOD) guidelines [26].

### 2.1. Source of data

This study was based on retrospective review of a prospective clinical database from a single Neurosurgery Department at the Walton Centre NHS Foundation Trust (UK). The Walton Centre contributes data to the Eurospine’s international Spine Tango registry [27], which governs standardised data collection protocols. Specifically, patients complete a self-assessment form (Core Outcome Measures Index, COMI) [28,29] at the preoperative consultation and postoperative follow-ups at 3, 12, and 24 months. The surgeon completes the surgery form before discharge, detailing the patient’s history, type of pathology and surgery, and hospital stay [30]. The Walton Centre’s database was reviewed on 26/04/2021 to extract lumbar spine surgery cases with degenerative disease as the main pathology. This data included patients operated between 4/04/2011 and 30/03/2021, with the last follow-up dated 21/04/2021.

### 2.2. Participants

Eligible patients had lumbar disc herniation and/or stenosis and underwent elective spinal decompression surgery with or without fusion. These most common degenerative pathologies and surgical measures were selected to obtain a representative sample and minimise clinical heterogeneity. Eligible patients completed preoperative and at least one postoperative self-assessment form. If there were multiple surgery cases per patient, only chronologically first eligible surgery was included.

### 2.3. Outcomes

The spinal surgery outcomes were defined as reduction in back and leg pain intensity and reduction in COMI from baseline to the last available follow-up. Due to the self-reported nature of outcomes, their assessment was not blinded.

Back and leg average pain intensity in the past week was measured on 0 (*no pain*) to 10 (*worst pain I can imagine*) numerical rating scales embedded within COMI. This scale is a recommended outcome measure of pain in studies evaluating effectiveness of treatments for chronic pain [31]. COMI [28] is a multidimensional instrument consisting of measures of pain, function, symptom-specific well-being, quality of life, social disability, and work disability. Average COMI score can range from 0 to 10, where higher scores indicate worse level of functioning. This instrument is an official outcome measure of Eurospine’s international spine registry and has been extensively validated in patients with back pain [29,32].

Both continuous and dichotomous outcomes were considered for their precision and clinical utility, respectively. Indeed, relative to dichotomisation, continuous outcomes provide greater statistical power, minimise information loss and risk of false positive results, and allow more accurate estimation of variability in outcomes (e.g. those close to and far from the MCID cut-off) [33]. Here continuous outcomes represent numerical differences between the baseline and follow-up back pain, leg pain, and COMI. Positive scores correspond to improvement (i.e., reduction in pain and functional impairment). Dichotomous outcomes represent achievement of MCID between baseline and follow-up back pain and leg pain (reduction of ≥2 points [34]), and COMI (reduction of ≥2.2 points [35]).

### 2.4. Predictors

As candidate predictors we assessed preoperative factors, and controlled for potential intra- and postoperative confounders (see **Table 1** for a full list). Assessment of all predictors was blinded to outcomes assessed ≥3 months later, but not blinded to other predictors, either self-reported by the patients before surgery, or recorded by the surgeon before discharge.

**Table 1.**
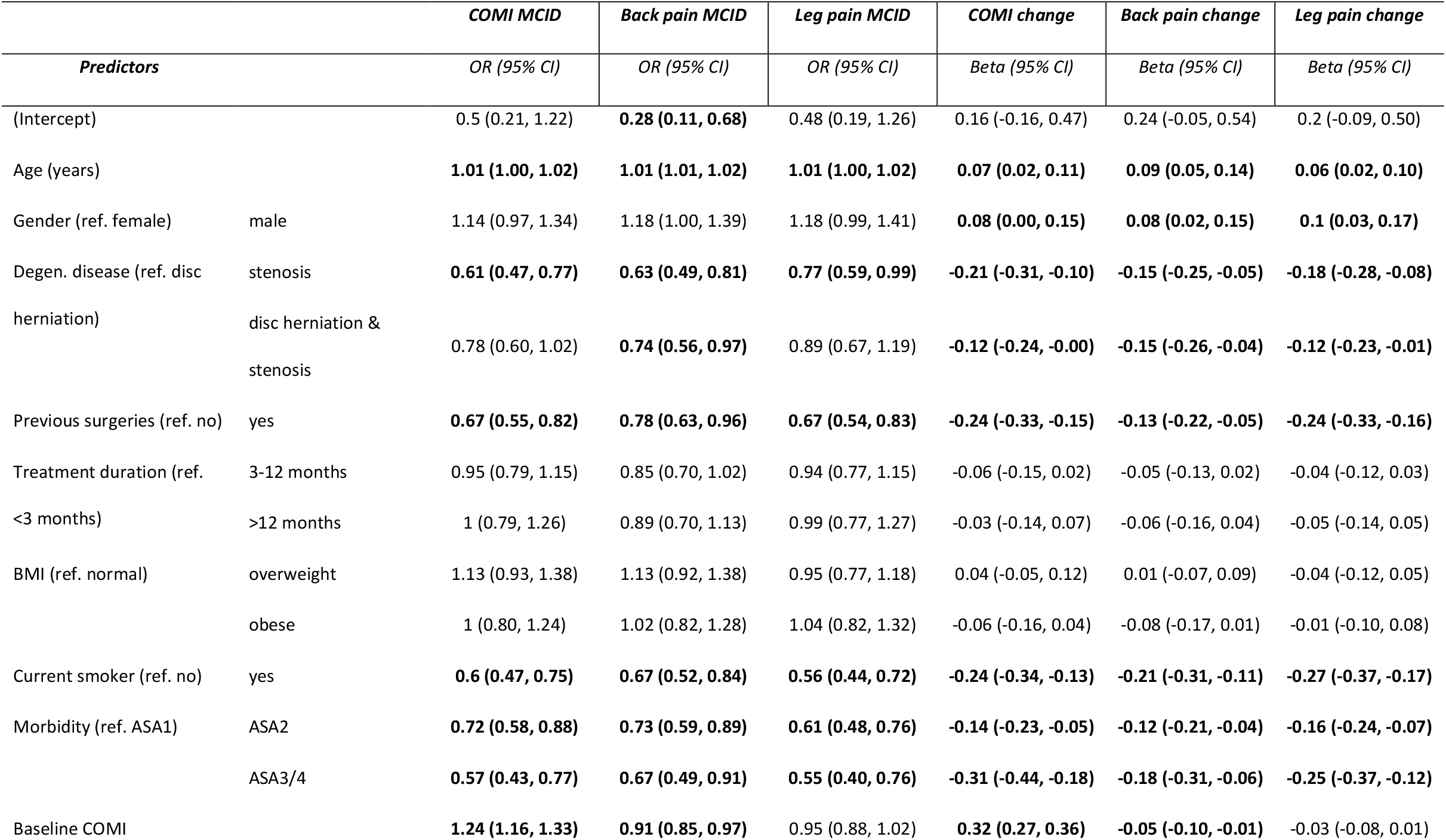
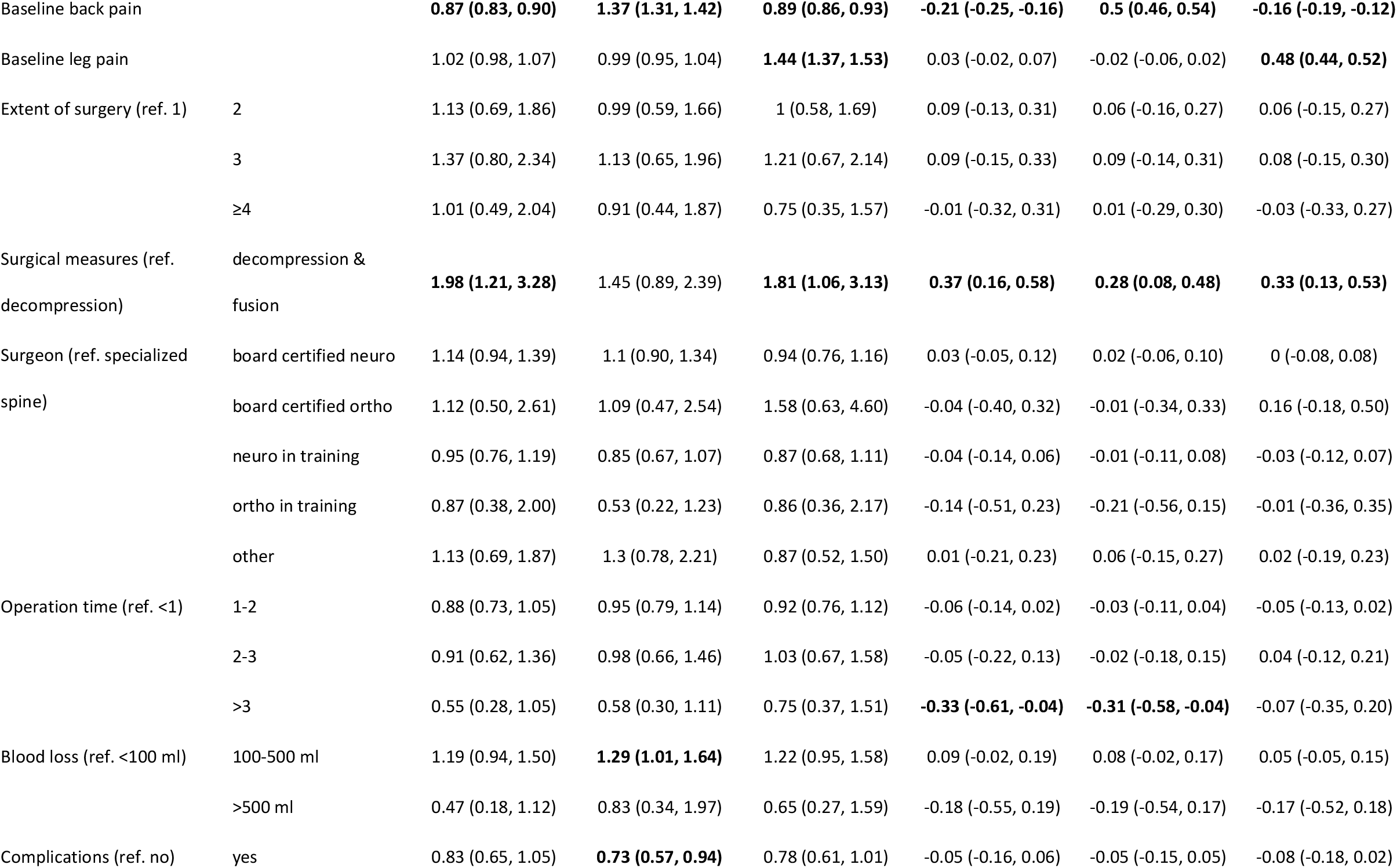
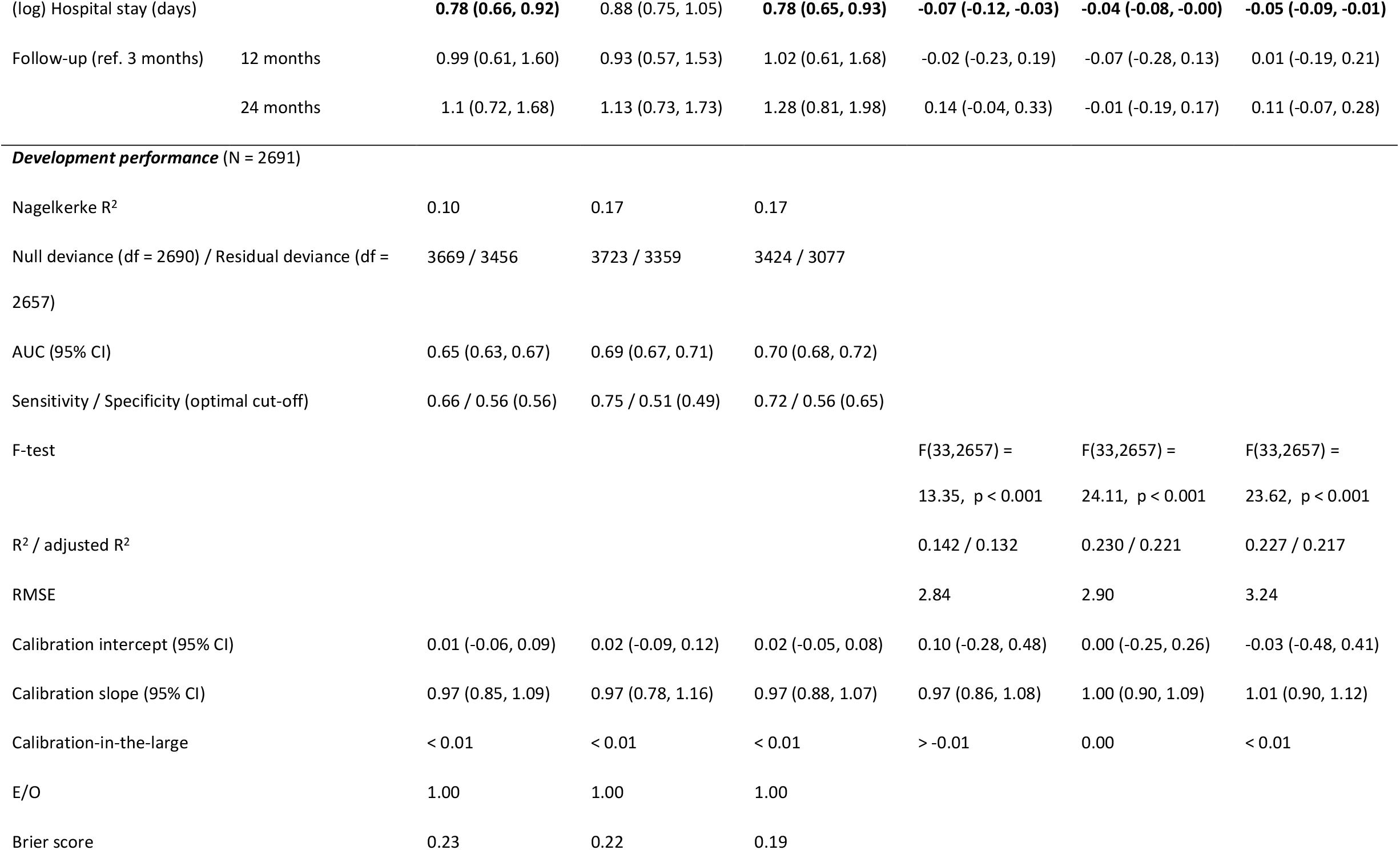
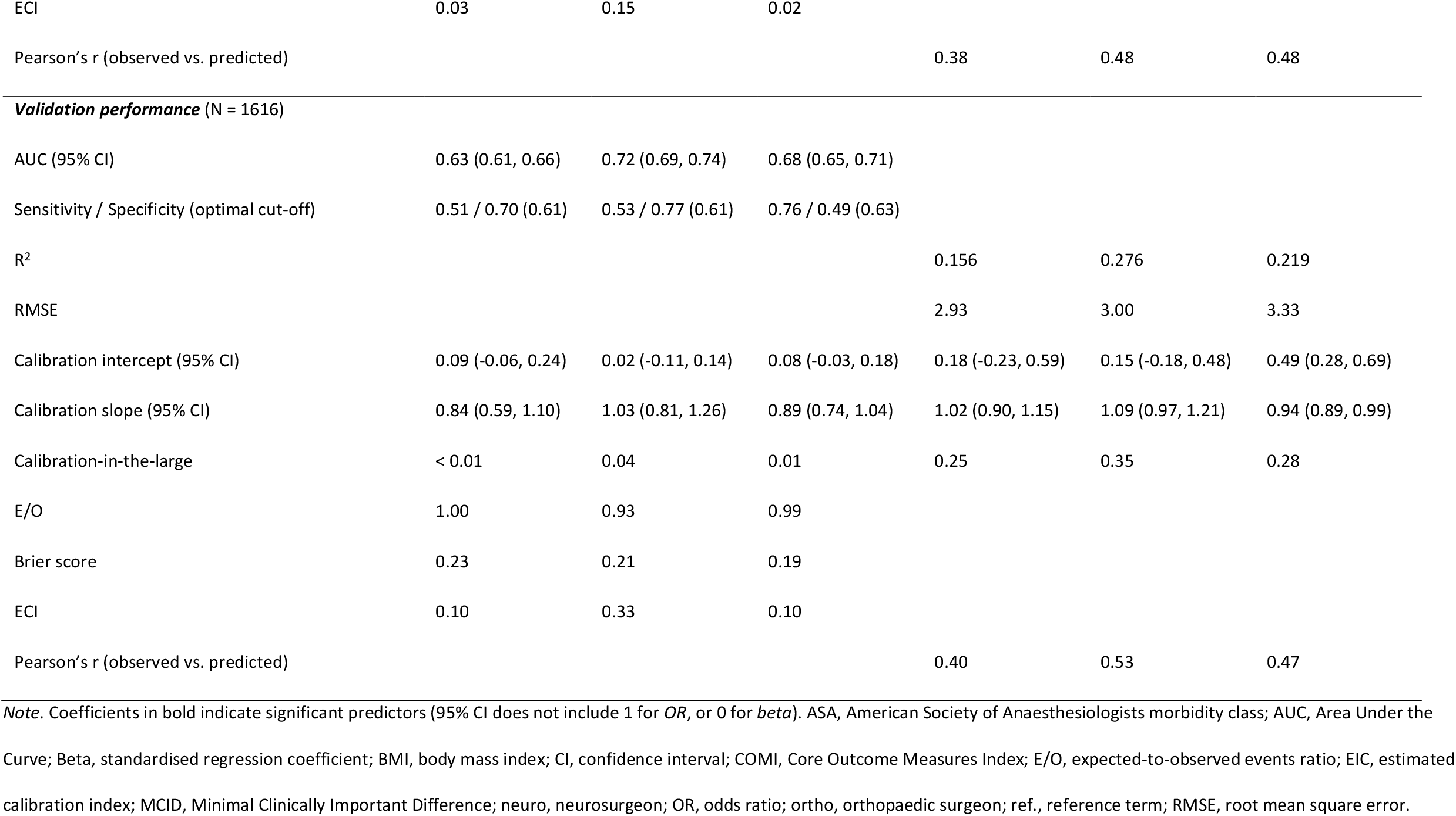
Multivariate regression models results in the development data and models’ performance in the validation data.

### 2.5. Sample size

To estimate the minimum required sample size and minimise the risk of overfitting, at least 20 participants per predictor should be included for continuous outcomes [36], and 10 or more events (i.e., achieving MCID) per predictor for dichotomous outcomes [37]. We considered 19 predictors, however, since several categorical factors had more than two levels, each additional factor level was counted separately, resulting in 34 predictor parameters in total. Therefore, a minimum of 680 participants would be required for continuous outcomes, and 340 participants who achieve MCID for dichotomous outcomes.

### 2.6. Data processing and statistical analysis

Data was processed and analysed using R software, version 4.1.1 [38]. Details of data processing and handling of predictors are described in **Methods S1, Additional file 1**. Except for categorising portion of the BMI data to match the variable types across two versions of the surgery form, other continuous factors were treated as continuous in the analysis. Due to severely skewed distribution, hospital duration was log-transformed (base 10).

#### 2.6.1. Handling missing data

As per our eligibility criteria, only patients with complete outcome data were included in the analysis. We report attrition and summary statistics on the portion of excluded data, and between-group comparisons with the final included sample.

Missing predictor data was assumed to be missing at random and addressed through Multivariate Imputation by Chained Equations (MICE) with 40 iterations using *mice* R package, [39]. The MICE procedure is explained in **Methods S2, Additional file 1**. Values were missing in 11 out of 19 predictors, with partial missing rates from <1% to 36% (see Results for details), and a total missing rate of 3.75% across all predictors. Multiple imputation has been found to remain unbiased up to 50% missing rates [40,41]. A series of diagnostic checks detailed in **Methods S2, Additional file 1** demonstrated good convergence and no apparent biases of the MICE algorithm, with overlapping range and distributions of the imputed and observed data (**Figs. S1-S4, Additional file 1**), and the missingness of each variable was associated with other factors in the imputation dataset (**Table S1, Additional file 1**), therefore supporting the missing at random assumption. We also conducted a sensitivity complete case analysis excluding cases with missing data on any of the predictors.

#### 2.6.2. Model development and validation data

The dataset was split into model development and validation samples based on the date of surgery (2011-2017 and 2018-2021, respectively). For large datasets, a non-random split has been recommended, e.g., by time [37]. The chosen time-split coincided with the introduction of a new version of the surgery form. The setting, eligibility criteria, predictors, and outcomes used were consistent across the development and validation data, except for some differences between the surgery form versions (see **Methods S1, Additional file 1**).

#### 2.6.3. Multivariate regression analysis

As our primary analysis, we developed multivariate logistic regression models for each dichotomous outcome, and multivariate linear regression models for each continuous outcome. Models were fitted using R functions *glm* (with binomial family and logit link function) and *lm* (for linear regression) within the *stats* package [38]. Full model approach (i.e., including all candidate predictors) was used to estimate prediction accuracy based on the available set of routinely collected data in combination and to assess relative contribution of specific preoperative factors while controlling for potential confounders. Each model based on the development sample was then fitted to the predictor values in the validation data to predict the outcomes of interest in new, untrained data. The predicted outcomes were compared with the actual (observed) outcome values.

Model performance on the development and validation data was expressed by calibration and discrimination measures with 95% confidence intervals (CI) where applicable. Note that although several internal validation methods are available, such as bootstrapping [42], our primary focus was on the validation on an external sample, which is a preferred, more robust, method to those based on resampling the internal data [37]. For calibration plots, we divided each dataset into 10 deciles according to the predicted probability of MCID (or predicted change score for linear regression), and plotted the mean predicted versus mean observed probabilities (or change scores) for each decile [43]. Perfect fit would be reflected by all points being aligned on a line with intercept 0 and slope 1. We tested whether the 95% CIs of the intercept and slope of the calibration line of best fit include these values. While this method is applicable to both dichotomous and continuous outcomes, other approaches have been proposed as more robust for assessing calibration of outcome probabilities, such as flexible calibration curves [44], which we present in **Additional file 1** for reference. We additionally calculated calibration-in-the-large (mean observed – mean predicted outcome probabilities or values), and for MCID models, reported expected-to-observed events ratio (E/O; 1 would indicate perfect calibration), Brier score (mean squared error between the observed and predicted outcome probabilities; 0 would indicate perfect calibration), and estimated calibration index (ECI; average squared difference between predicted and observed outcome probabilities transformed into a single number [0-1] summarising a flexible calibration curve, with 0 indicating perfect calibration) [44,45]. For linear regression models, also a correlation between ungrouped observed and predicted outcomes was expressed as Pearson’s *r*. Discrimination of the logistic regression models was assessed via Receiver-Operating Characteristic curve (ROC) plots with estimated Area Under the Curve (AUC; c-index), and classification accuracy expressed as sensitivity and specificity estimates at the optimal ROC cut-off (probability threshold maximising both indices). AUC can range from 0 to 1 and a value of 0.5 corresponds to chance discrimination, while 0.7-0.8 is considered acceptable, and >0.8 excellent discrimination [46]. Nagelkerke pseudo-*R*^*2*^ and deviance were also reported as overall performance measures. For linear regression, discrimination was quantified by *R*^*2*^, *F*-test, and root mean square error (RMSE) for each model. To express adjusted contribution of each predictor to the outcome of interest, we presented log odds and odds ratios (for dichotomous outcomes) and unstandardised and standardised regression coefficients (for continuous outcomes) with 95% CIs.

#### 2.6.4. Random forests

As our secondary analysis, we used random forests (RF), as a particularly flexible machine learning approach, which can combine different data types and be applied to both dichotomous and continuous outcomes, and in some studies demonstrated superior predictive performance over other machine learning algorithms [47–49]. RF is a non-linear classification and regression algorithm based on an ensemble of deep decision trees. Multiple decision trees are trained on different randomly selected bootstrap samples of the same training dataset. Each time a decision tree split is performed, the best split variable [47] is chosen from a random subset of the original predictor set. Each tree gives an outcome prediction on the leftover data which was not used during training (out-of-bag, OOB). The predictions are then aggregated (bagging) by assigning class labels (MCID vs. no-MCID) by majority vote, or by averaging the continuous dependent variable (change score) across all the trees. Prediction errors are calculated as OOB error (misclassification rate) for classification or OOB mean square error for regression. We used Breiman’s implementation of RF in *randomForest* R package [47,50]. The RF algorithm hyperparameters were tuned to maximise the predictive accuracy as described in **Methods S3** and **Table S2, Additional file 1**. Using the final hyperparameters and 500 trees for each model, we applied RF classification to predict MCID outcomes, and RF regression to predict continuous outcomes in the development data. Then we used each developed model to predict the same outcomes in the validation data. Although the leg pain outcome was characterised by a moderate class imbalance, with MCID rates of 67-68%, no adjustments such as downsampling non-events were made, as these could distort the true outcome rates, lead to inadequate clinical predictions, and increase the risk of overfitting.

RF discrimination in the development and validation data was quantified by OOB errors and AUC with sensitivity and specificity at the optimal ROC cut-off for classification models; and by RMSE, pseudo-*R*^*2*^, and Pearson’s *r* for observed against predicted outcomes for regression models. Calibration plots were prepared following the procedures described for the logistic and linear regression models. Calibration was additionally quantified by calibration-in-the-large, and for classification models also E/O, Brier score, and ECI with supplemental flexible calibration curves (**Additional file 1**). We also presented relative variable importance based on mean decrease in accuracy (loss in prediction performance) when a particular variable is omitted from the training data for each development model.

## 3. Results

### 3.1. Participants

**Fig. S5, Additional file 1** illustrates the flow of participants through the eligibility screening process. Out of 6810 screened surgery cases, 4307 unique patients were included in the analysis. Descriptive characteristics, missingness rates, and statistical comparisons between included and excluded participants due to missing baseline and/or follow-up assessment are reported in **Results S1** and **Table S3, Additional file 1**. Although there were statistically significant differences on 10 predictors, as expected in large datasets even when effect sizes of these differences are small, the included data were representative: the included data covered the full range of possible predictor values in the excluded data, and all levels of categorical factors present in the excluded data were well-represented in the included data.

Development dataset included 2691 and validation dataset 1616 patients, thus each was more than sufficient to fit regression models with 34 specified predictor parameters. Patient characteristics are presented in **Table S4**, and any group differences and additional post-hoc sample size considerations based on the observed outcome rates and means are described in **Results S1, Additional file 1**. On average, patients in the development compared to the validation sample achieved less reduction in back pain (mean [SD]: 2.15 [3.30] vs. 2.51 [3.50]) and leg pain (3.68 [3.69] vs. 3.94 [3.75]), but did not differ in COMI change (3.22 [3.07] vs. 3.34 [3.18]) or MCID rates in COMI (57% vs. 58%), back pain (53% vs. 57%), or leg pain (67% vs. 68%). There were slight statistically significant differences on most of the candidate predictors; yet, importantly, the validation data was within the range of the development data for all variables.

### 3.2. Development and validation of regression models

Regression diagnostic checks are detailed in **Results S2** and **Figs. S7-11, Additional file 1**. In summary, there was no multicollinearity among the predictors, no severe deviations from linearity between the continuous predictors and logit of MCID or continuous outcomes, residual variance was homogenous across different levels of categorical predictors, there were no highly influential values, models had good fit across the range of observations, and standardised residuals in linear models showed acceptable homoscedasticity and normal distribution, with slight deviations on the tails.

#### 3.2.1. Binary outcomes (MCID)

**Table 1** presents odds ratios with 95% CIs for each candidate predictor in the development models for achievement of MCID in COMI, back pain, and leg pain intensity (see also **Fig. S12, Additional file 1**). Independent of other factors included in the models, older age was associated with higher odds of achieving MCID after surgery across all outcomes. Decompression with fusion surgery was related to higher odds of MCID in COMI and leg pain, whereas blood loss of 100-500 ml – higher odds of MCID in back pain. Additionally, higher baseline COMI, back pain, and leg pain predicted better odds of improvement in their corresponding outcomes. In contrast, patients with spinal stenosis, history of previous surgeries, currently smoking, and with higher morbidity class had lower odds of achieving MCID after surgery across all outcomes. Disc herniation with stenosis, higher baseline COMI, and presence of any complications also predicted lower odds of MCID in back pain. Finally, patients with higher baseline back pain and longer hospital stay had lower odds of MCID in COMI and leg pain. Adjusted effect sizes of these predictors were small (odds ratios >0.4 and <2.5).

The AUC in the development and validation data was bordering on between no-better-than-chance and acceptable discriminability (**Table 1**). COMI MCID model had the worst discrimination, whereas the highest, acceptable, discrimination ability was found for the back pain MCID model in the validation data. Using the optimal ROC cut-offs for each outcome, development models generally had good sensitivity (ability to detect true MCID), while specificity (detecting true no-MCID) oscillated near chance classification. There was a consistent pattern of classification in the validation data for leg pain MCID, however, COMI and back pain MCID classification presented an opposite pattern, with good specificity but poor sensitivity. The ROC curves are presented in **Fig. S13a, Additional file 1** for development data, and **Fig. 1a** for validation data.

**Fig. 1.**
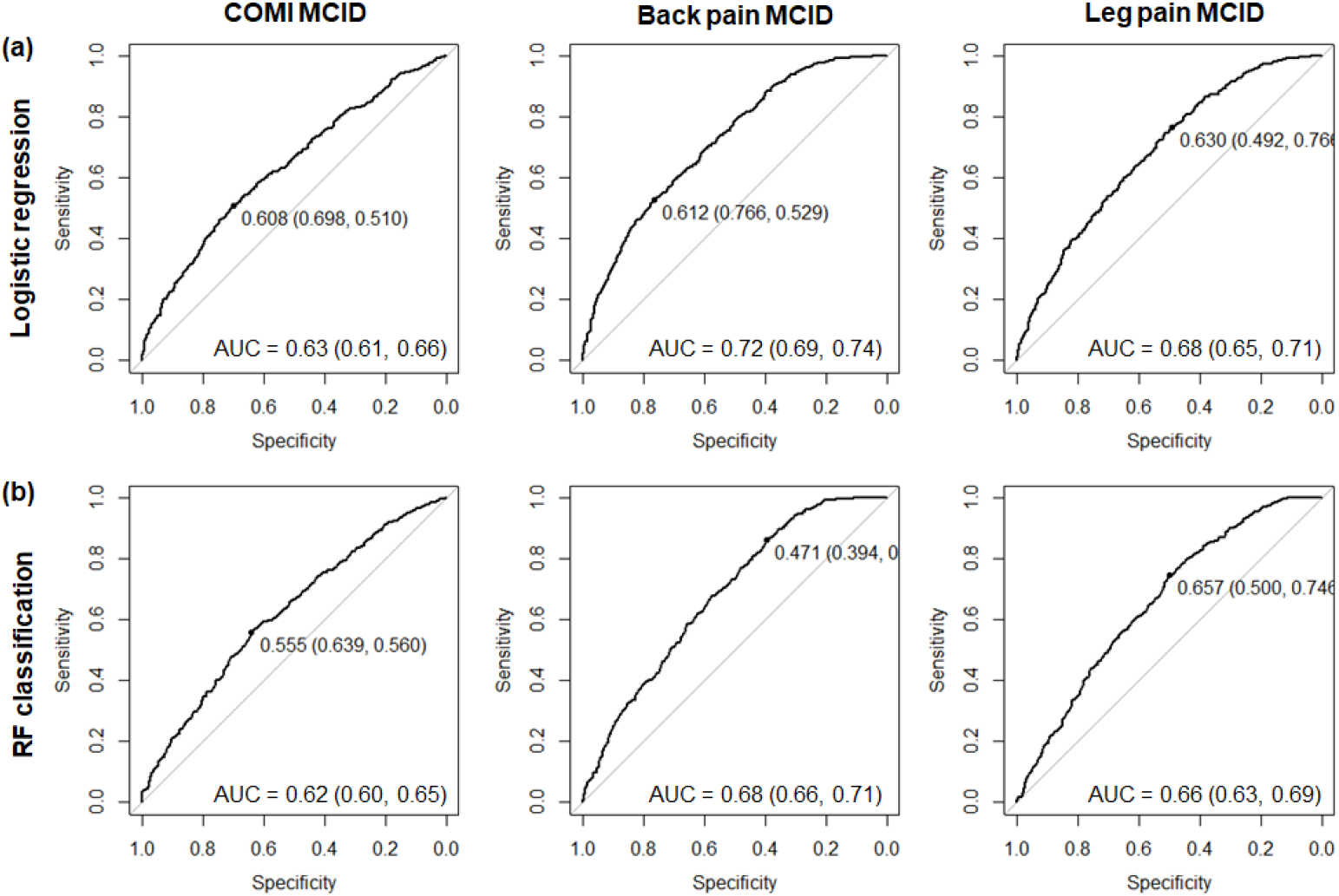
Discrimination ability of the (a) logistic regression and (b) random forest classification models when fitted to the validation data for MCID in COMI, back pain, and leg pain. Plots illustrate Receiver-Operating Characteristic (ROC) curves with an optimal probability threshold (black point on the ROC curve; specificity and sensitivity indicated in brackets). Area Under the Curve (AUC) is reported for each ROC with 95% confidence interval.

The proportion of explained variation in the development models ranged from 10% for COMI to 17% for back and leg pain intensity. Residual deviance was lower than null deviance, indicating that the included variables allow to predict each outcome better than intercept-only (null) models. Calibration-in-the-large was near zero and E/O equal to or approaching 1, suggesting no overall differences between mean observed and predicted outcomes. Brier scores ranged from 0.19 to 023, consistently across the development and validation models (**Table 1**). Calibration plots indicated good model fit for MCID outcomes in the development data (**Fig. S14a, Additional file 1**). In the validation data, considering the visual inspection (**Fig. 2a**) and the fact that in all cases, the intercept of the calibration lines did not significantly differ from 0, and their slope did not significantly differ from 1, we conclude that the models showed good external calibration. On a more granular level, flexible calibration curves for COMI and leg pain MCID consistently showed good calibration, whereas that for the back pain model, accompanied by a higher ECI, had a positive intercept and suggested a small degree of underestimation, particularly in the range of 0.3-0.5 predicted MCID probabilities (**Fig. S15a, Additional file 1**).

**Fig. 2.**
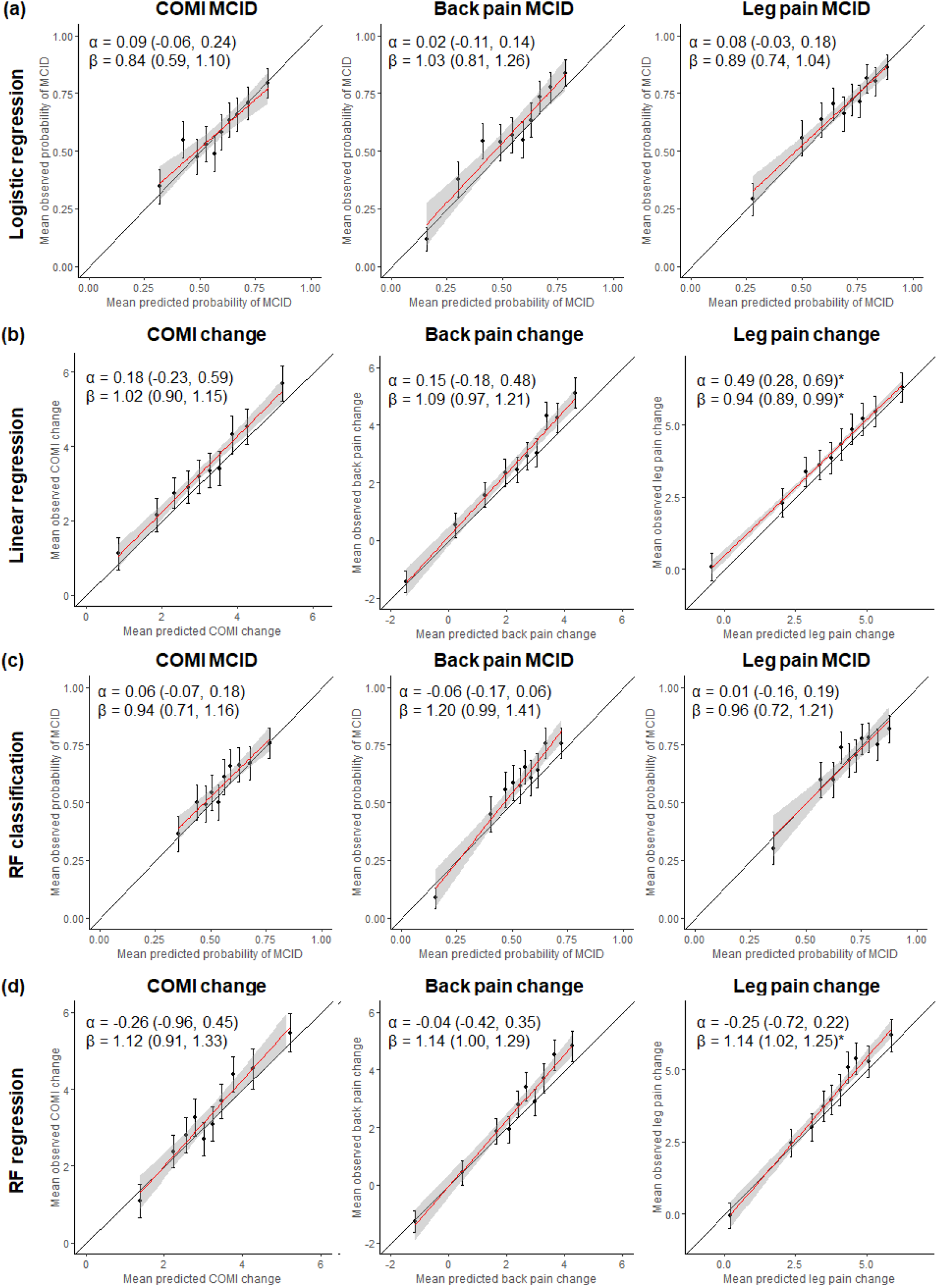
Calibration plots for (a) logistic regression, (b) linear regression, (c) random forest (RF) classification, and (d) RF regression models when fitted to the validation data for each outcome. Points correspond to the mean predicted and observed probabilities of MCID or change scores in each decile with 95% confidence intervals (CI) of the mean observed probabilities or change scores. Calibration lines of best fit are plotted in red in 95% CI in grey and their intercept (α) and slope (β) estimates with 95% CIs are presented in the top-left corner of each plot. *95% CI of the intercept do not include 0, or the 95% CI of the slope do not 1, indicating significant deviation from the perfect fit.

#### 3.2.2. Continuous outcomes

Results of the linear regression models on change in COMI, back pain, and leg pain are presented in **Table 1** (standardised regression coefficients) and **Fig. S12, Additional file 1**. After adjusting for other factors included in the models, older age, male gender, and decompression with fusion (moderate effect size) were associated with greater improvement after surgery across all outcomes. Additionally, higher baseline COMI, back pain, and leg pain predicted greater improvement in their corresponding outcomes (moderate-large effects). On the contrary, patients with spinal stenosis and disc herniation with stenosis, history of previous surgeries, currently smoking (moderate effect), with higher morbidity class (moderate effect), and longer hospital stay had less improvement after surgery across all outcomes. Additionally, higher baseline COMI predicted less improvement in back and leg pain, higher baseline back pain predicted less improvement in COMI and leg pain, and operation time >3 hours (moderate effect) predicted less improvement in COMI and back pain. Adjusted effect sizes of these predictors were small (*betas* <0.25), unless specified otherwise.

Discrimination performance in the development models ranged from 13% (COMI change) to 22% (back and leg pain change) and the included set of predictors explained each outcome better than intercept-only models (*F*-test *ps* <0.001) (**Table 1**). Unadjusted portion of explained variance was higher in the validation than development data for COMI and back pain change, but lower for leg pain change outcome. Prediction accuracy was slightly worse in the validation compared to the development data with approximately 0.1 higher RMSEs across all outcomes.

Model calibration was very good in the development data (**Fig. S14b, Additional file 1**), and plots in the validation data also showed close agreement between mean observed and predicted outcomes, although there was some degree of underestimation of the predicted changes in back and leg pain (**Fig. 2b**), also apparent in flexible calibration curves (**Fig. S16a, Additional file 1**). However, only the leg pain model calibration line significantly deviated from the perfect fit. Calibration-in-the-large indicated that average predicted changes in COMI and pain outcomes were 0.25-0.35 points lower than observed changes (**Table 1**).

Individual predictions of outcomes based on the developed logistic and linear regression models can be made according to the equations provided in **Results S3** and log-odds and unstandardised regression coefficients presented in **Table S5, Additional file 1**.

Compared to the primary analyses on the imputed data, sensitivity complete case analyses presented in **Results S4** and **Table S6** showed similar or worse model performance in the development and validation datasets, and no systematic differences in significant predictors except for the current smoking status which did not significantly predict any outcomes in the sensitivity analyses.

### 3.3. Development and validation of random forest models

**Table 2** provides an overview of the RF performance measures across MCID (classification) and continuous change (regression) in COMI, back pain, and leg pain outcomes in the development and validation data.

**Table 2.** Random forests performance measures in the development and validation data across all outcomes.

|  | *Classification outcomes* |  |  | *Regression outcomes* |  |  |
| --- | --- | --- | --- | --- | --- | --- |
|  | *COMI MCID* | *Back pain MCID* | *Leg pain MCID* | *COMI change* | *Back pain change* | *Leg pain change* |
| **Development** |  |  |  |  |  |  |
| AUC (95% CI) | 0.62 (0.60, 0.64) | 0.66 (0.64, 0.68) | 0.67 (0.64, 0.69) |  |  |  |
| Sensitivity / Specificity (optimal cut-off) | 0.51 / 0.68 (0.58) | 0.67 / 0.56 (0.53) | 0.71 / 0.53 (0.67) |  |  |  |
| OOB error / RMSE | 39.84% | 38.72% | 29.77% | 2.89 | 2.94 | 3.33 |
| pseudo-R<sup>2</sup> |  |  |  | 0.112 | 0.207 | 0.186 |
| Calibration intercept (95% CI) | 0.11 (0.04, 0.18) | 0.01 (-0.08, 0.10) | 0.01 (-0.12, 0.14) | 0.04 (-0.46, 0.54) | -0.06 (-0.33, 0.22) | -0.18 (-0.64, 0.28) |
| Calibration slope (95% CI) | 0.83 (0.71, 0.95) | 1.00 (0.83, 1.16) | 0.95 (0.76, 1.13) | 0.98 (0.84, 1.13) | 1.02 (0.92, 1.13) | 1.05 (0.93, 1.17) |
| Calibration-in-the-large | 0.01 | < 0.01 | -0.03 | -0.02 | -0.01 | < 0.01 |
| E/O | 0.98 | 0.99 | 1.04 |  |  |  |
| Brier score | 0.23 | 0.22 | 0.20 |  |  |  |
| ECI | 0.07 | 0.09 | 0.16 |  |  |  |
| Pearson's r (observed vs. predicted) |  |  |  | 0.33 | 0.45 | 0.43 |
| **Validation** |  |  |  |  |  |  |
| AUC (95% CI) | 0.62 (0.60, 0.65) | 0.68 (0.66, 0.71) | 0.66 (0.63, 0.69) |  |  |  |
| Sensitivity / Specificity (optimal cut-off) | 0.56 / 0.64 (0.56) | 0.86 / 0.39 (0.47) | 0.75 / 0.50 (0.66) |  |  |  |
| RMSE |  |  |  | 2.94 | 3.04 | 3.33 |
| pseudo-R<sup>2</sup> |  |  |  | 0.152 | 0.252 | 0.220 |
| Calibration intercept (95% CI) | 0.06 (-0.07, 0.18) | -0.06 (-0.17, 0.06) | 0.01 (-0.16, 0.19) | -0.26 (-0.96, 0.45) | -0.04 (-0.42, 0.35) | -0.25 (-0.72, 0.22) |
| Calibration slope (95% CI) | 0.94 (0.71, 1.16) | 1.20 (0.99, 1.41) | 0.96 (0.72, 1.21) | 1.12 (0.91, 1.33) | 1.14 (1.00, 1.29) | 1.14 (1.02, 1.25) |
| Calibration-in-the-large | 0.02 | 0.05 | -0.01 | 0.14 | 0.28 | 0.25 |
| E/O | 0.96 | 0.92 | 1.02 |  |  |  |
| Brier score | 0.23 | 0.22 | 0.20 |  |  |  |
| ECI | 0.07 | 0.38 | 0.17 |  |  |  |
| Pearson's r (observed vs. predicted) |  |  |  | 0.39 | 0.5 | 0.47 |

AUC, Area Under the Receiver-Operating Characteristic Curve; CI, confidence interval; E/O, expected-to-observed events ratio; ECI, estimated calibration index; MCID, Minimal Clinically
Important Difference; OOB error, out-of-bag misclassification rate; RMSE, root mean square error.

#### 3.3.1. Binary outcomes (MCID)

Misclassification rates ranged from 30% for leg pain to 40% for back pain and COMI MCID outcomes. Discrimination performance was in a similar range for development and validation data, with AUC consistently over 0.60, but still below acceptable discrimination across all outcomes, and consistently lower than AUC values obtained from logistic regression models. Outcome classification at the optimal probability threshold in the development and validation data across back and leg pain outcomes appeared to be biased towards higher sensitivity at the expense of specificity which oscillated near or below chance classification of no-MCID cases. This pattern was reversed for COMI outcome, where classification showed better specificity but near-chance sensitivity. The ROC curves are presented in **Fig. S13b, Additional file 1** for development data, and **Fig. 1b** for validation data.

Calibration plots demonstrated good agreement between mean observed and predicted MCID probabilities in the development data, however, the calibration line for the COMI outcome significantly deviated from the perfect fit suggesting some underestimation of the predicted MCID (**Fig. S14c**). Calibration of the COMI and back pain models in the validation data also suggested a small degree of underestimation of predicted outcome probabilities, but without significant deviations from the perfect fit (**Fig. 2c**). However, higher-resolution flexible calibration curve for back pain MCID with a positive intercept and slope >1 further suggested some degree of underestimation of predicted probabilities in the 0.4-0.6 range of the validation data (**Fig. S15b, Additional file 1**). Back and leg pain calibration curves were also accompanied by higher ECIs compared to the logistic regression models. Nonetheless, RF models were characterised by comparable calibration-in-the-large, E/O, and Brier scores, and calibration plots supported good agreement between the mean observed and predicted probabilities of MCID for most quantiles in the validation data, similar to the calibration of the logistic regression models.

#### 3.3.2. Continuous outcomes

The proportion of explained variance in change in COMI, back pain and leg pain ranged from 11% for COMI to 21% for back pain in the development data, and increased in the validation data (15-25%). These values were lower compared to R^2^ from the linear regression models in the development data, but did not differ in the validation data except for the back pain model. RMSEs were only minimally higher in the RF regression, but with less discrepancy between the development and validation data.

RF regression models showed very good calibration in the development data (**Fig. S14d**). While there was also good agreement between mean observed and predicted outcomes in the validation data, calibration plots showed some degree of underestimation of the predicted relative to the observed changes in outcomes, in particular in the higher deciles, with the slope of the leg pain calibration line significantly deviating from the perfect fit (**Fig. 2d**). Similar trends are apparent in the flexible calibration curves (**Fig. S16b, Additional file 1**). Tendency to underestimate predicted changes in the validation data was similar to that in the calibration of linear regression models, although underestimation of leg pain change was more pronounced in lower deciles. Correlation coefficients between ungrouped observed and predicted outcomes in the RF models were overall higher in the validation than development data, and marginally lower compared to linear regression models. Calibration-in-the-large indicated smaller (relative to linear regression models) differences between average observed and predicted outcomes, in the range of 0.14-0.28 points.

#### 3.3.3. Variable importance

Highest variable importance in RFs was generally assigned to the baseline scores on the corresponding outcome measures (except for COMI MCID), for instance, baseline back pain was most important for classifying back pain MCID and predicting change in back pain intensity (**Fig. S17, Additional file 1**). While for pain intensity outcomes, these baseline scores appeared to be the sole most relevant predictors, COMI outcomes showed broader distribution of importance over different predictors. Across all outcomes, relatively high importance was also attributed to the duration of hospital stay, age, baseline scores on other outcome measures, current smoking status, type of degenerative disease, history of previous spinal surgeries, and morbidity. The same factors were found to have significant prognostic effects in the logistic and linear regression models.

## 4. Discussion

We developed and externally validated multivariate regression and RF models to predict patient-reported outcomes 3-24 months after lumbar spine surgery based on prospectively recorded medical and patient data. The models demonstrated good calibration in the temporal validation data, while their discrimination ability oscillated between acceptable and no-better-than-chance. Linear and logistic regression models performed better than RF algorithms, both in the development and validation data. The most important predictors included age, baseline COMI and pain scores, type of degenerative disease, previous surgeries, smoking, morbidity, and hospital stay.

This study brings a novel contribution to the field by assessing and comparing performance of linear and logistic regression models versus RF regression and classification algorithms, and validating them on external data, in order to further develop our ability to predict more precisely individual surgery outcomes. High number of participants and events per variable, which were limited in previous clinical prediction models [48], add to the strength of the present work. Comparable performance and consistency in identified predictors demonstrate the robustness and generalisability of our models across different patient-reported outcomes (COMI, back, and leg pain MCID and continuous change scores) and modelling approaches.

### 4.1. Model performance

There was no substantial decrease in the models’ performance on the new data relative to the development data, indicating no overfitting issues. Regression models predicting changes in back pain showed the best external validity, with acceptable discrimination (0.72) and 28% explained variance, followed by the models predicting leg pain outcomes (AUC 0.68, 22% explained variance). These metrics are comparable or better than in similar externally validated models predicting pain-related outcomes (AUC 0.52-0.83, 6-19% [17–19]). COMI models showed poorer discrimination (AUC 0.63, 16% explained variance), although comparable with external validity of another model relying on the same measure (17% [18]), suggesting that composite outcomes like COMI may be more difficult to predict than, for instance, specific disability measures (AUC 0.71 [19]).

Similar studies relying on internal validation generally reported better discrimination (AUC 0.64-0.84, 23-49% [12–16]), highlighting a potential degree of over-optimism when model performance is only assessed on resampled or randomly-split data. Furthermore, model calibration was not always assessed [12,16,17], but accurate prediction of outcomes can be particularly problematic in external data [18,19]. Our models showed good calibration in the validation data across all outcomes, although there was a mild tendency to underestimate back pain MCID and leg pain reduction.

We found that RF did not outperform linear and logistic regression models. RF showed similarly good calibration in the validation data, yet none of the models reached acceptable discriminability in the validation (or development) data (0.62-0.68). This may reflect the difficulty of RF algorithms to extrapolate to new, untrained data, although previous relevant studies only achieved RF discrimination of 0.64-0.72 in internal validation [21,22]. Various machine learning classification approaches (e.g. elastic net penalised regression, deep neural networks, extreme gradient boosting, RF) have previously shown superior predictive performance compared to logistic regression [20,22,23]. However, consistent with our findings, a recent meta-analysis concluded that based on low risk of bias studies, performance of machine learning clinical prediction algorithms, including RF, does not differ from logistic regression (their advantage was only found in high risk of bias studies) [48]. Our results extend this conclusion to linear regression versus RF regression.

There could be several reasons why RFs did not outperform statistical regression in the present study. Previous work demonstrating an advantage of machine learning over logistic regression did not cover external validation [20,22,23], while regression models are likely to have better generalisability. Furthermore, machine learning works best for problems with high signal-to-noise ratio, which rarely characterises clinical data. Finally, since RFs show improved performance on data with nonlinear and nonadditive effects, any nonlinearities in the present data were likely not severe enough to be detrimental to statistical regression. Therefore, RFs might not show superior performance on large enough datasets satisfying the regression assumptions.

### 4.2. Relevant predictors

According to the regression models, greater odds of achieving MCID and larger reduction in COMI, back, and leg pain were significantly associated with older age, higher baseline score on the respective outcome measure, having decompression surgery with fusion, no stenosis, no history of previous spinal surgeries, lower morbidity class, not smoking, and shorter hospital stay. The same factors (except for surgical measures) were the most important predictors in RF analyses, with preoperative COMI, back, or leg pain scores leading across all models. Relevance of several of the identified predictors was also supported by previous research on patient-reported outcomes from the Spine Tango registry in other countries [18,51]. Our results are also consistent with systematic reviews supporting prognostic value of age, preoperative pain intensity and disability, type of spinal pathology, previous surgeries, and smoking [6–10]. In contrast, we did not find any effect of symptom duration, here recorded indirectly as duration of previous treatment.

Low back pain and spinal surgery are complex clinical issues where multifactorial data is necessary to make accurate individualised predictions of treatment outcomes. Suboptimal model performance, particularly on COMI and leg pain outcomes, suggests that additional factors to those already recorded in registries such as Spine Tango are likely needed to improve the predictive accuracy. For instance, other predictors identified in the above-mentioned systematic reviews, but not available in our data, included education level, compensation, duration of sick leave, sensory loss, comorbidities, and psychological pain-related and affective factors. Previous prediction models which achieved better discrimination (at least in internal validation) incorporated additional predictors, such as unemployment, medical insurance (although not applicable in the UK context), opioid use, antidepressants, mental functioning, optimism, control over pain, catastrophising, and postoperative psychomotor therapy [12–14,21].

### 4.3. Limitations

The present study is not without limitations. Class imbalance is a common problem for machine learning classification algorithms, where selecting the outcome occurring more frequently increases overall classification accuracy, which may still be poor for the less frequent outcome [52]. This could potentially account for the sensitivity/specificity trade-off apparent in some of the MCID models, although not specific to RF.

Furthermore, there was missing data on several predictors, and although imputation diagnostics did not indicate any biases, complete case sensitivity analysis was inconsistent with respect to the predictive value of smoking status, which had the highest missingness rate. This suggests that the imputed data may not accurately reflect the true smoking status in the population of interest. Thus, the missingness of the smoking status could potentially be related to other unmeasured variables. However, the significant prognostic value of smoking is consistent with several previous prediction models for spinal surgery outcomes [14,16–18].

Finally, although the developed models performed relatively well in the temporal validation, geographic validation is often more problematic [53]. Thus, future research could include external validation of the prediction models across different neurosurgery centres to further assess their generalisability.

### 4.4. Implications

The developed models demonstrated good ability to predict spinal surgery outcomes from new data, thus in practice, they could help identify patients at risk of poor outcomes. Such patients could be considered for additional interventions to improve their chance of recovery [12]. While all models appeared to be well-calibrated, and those predicting change in back pain showed the best performance on external validation, the discrimination ability of the leg pain and COMI models could be further improved, for instance, by including factors that previously demonstrated important contributions to spinal surgery outcomes. Modifiable preoperative predictors could be particularly useful for prospectively maximising the treatment benefit. The proposed models can therefore serve as a benchmark to inform future studies aimed at improving the accuracy of individual outcome prediction and potential revision of routinely collected information for spinal surgery registries.

## 5. Conclusions

We found comparable performance and consistent predictors across different outcomes, modelling approaches, and datasets. Regression models showed good calibration and acceptable to no-better-than-chance discrimination in the validation data. For similar datasets (with comparable set of predictors, sufficient sample size, and satisfying regression assumptions), RFs do not appear to outperform statistical regression. A strong advantage of statistical regression is its explanatory value and more easily interpretable prediction rules readily applicable in the clinical context. RFs, however, allow to establish relative predictor importance, which may assist in prioritising complex multifactorial data. Nonetheless, there is still room for improvement in terms of recorded predictor data.

## Data Availability

Due to confidentiality, the data are not publicly available.

## List of abbreviations

AUC: area under the curve
BMI: body mass index
CI: confidence interval
COMI: core outcome measures index
E/O: expected-to-observed events ratio
ECI: estimated calibration index
MCID: minimal clinically important difference
MICE: multivariate imputation by chained equations
OOB: out-of-bag
RF: random forest
RMSE: root mean square error
ROC: receiver-operating characteristic curve
TRIPOD: Transparent Reporting of a multivariable prediction model for Individual Prognosis Or Diagnosis

## Declarations

### Ethics approval and consent to participate

Patients partaking in the Spine Tango registry consented for their pseudonymised data being used for research purposes, however, due to confidentiality, the data are not publicly available. The study received approval from University of Liverpool Health and Life Sciences Research Ethics Committee (ref. 8224).

### Consent for publication

Not applicable.

### Availability of data and materials

The data that support the findings of this study are available from the Walton Centre NHS Foundation Trust but restrictions apply to the availability of these data, which were used under information sharing agreement for the current study, and so are not publicly available.

Materials such as analysis scripts used in the current study are available at https://osf.io/x3rwg/?view_only=958cac2415e54281980c83ff7f4d786c.

### Authors’ contributions

Conceptualization: CB, MW, RD, MH; Data Curation: MH; Formal Analysis: MH; Funding Acquisition: CB, MW, RD; Investigation: MH; Methodology: MH, CB; Project Administration: MH, CB; Resources: MH, MW; Software: MH; Supervision: CB; Validation: MH, CB; Visualization: MH; Writing – Original Draft Preparation: MH, CB; Writing – Review & Editing: MH, CB, MW, RD. All authors discussed the results and commented on the manuscript, and read and approved its final version.

## Acknowledgements

Not applicable.

