## Supplement for "Predicting patient-reported outcomes following lumbar spine surgery: development and external validation of multivariable prediction models"

### Additional file 1

#### Methods S1: Data processing and handling of predictors

We extracted total COMI scores, back pain ratings, and leg pain ratings from self-assessment form at baseline and each follow-up interval. Change scores for each outcome were calculated by subtracting the last available follow-up score from the baseline score. Achievement of MCID at the last available follow-up on each outcome was coded as TRUE if the change score was ≥2.2 for COMI, and ≥2 for pain intensity. Otherwise, it was coded as FALSE.

Due to change in the surgery form during the period covered by this study, there were some differences in how the variables of interest were recorded. We refer to the two form versions as 2011 and 2017.

(a) Duration of symptoms requiring treatment in 2017 version included fewer categories (<3, 3-12, >12 months) compared to the 2011 version (<3, 3-6, 6-12, >12 months), therefore, we combined '2-6 months' and '6-12 months' into a single '3-12 months' category. 2011 version also included a distinction between surgical and conservative treatment, however, since only conservative treatment had any duration specified, and history of previous spinal surgeries was recorded as a separate factor, surgical treatment category was disregarded.

(b) Presence of flags (each colour as a separate category) was only recorded in the 2011 version, therefore, this information was not included in our primary analyses.

(c) The 2017 version included a distinction between primary and secondary type of degeneration, which was not present in the 2011 version and not possible to determine retrospectively from the available data. Therefore, primary and secondary type of degeneration were combined into a single variable. From both surgery forms, we extracted the information whether disc herniation or central / lateral / foraminal stenosis were present and coded the categories as disc herniation (alone), stenosis (alone), and disc herniation with stenosis.

(d) Extent of surgery in the 2017 version was recorded separately for each surgical measure, whereas the 2011 version recorded one extent regardless of surgical measures. Therefore, for the 2017 version data, we combined the extent of the eligible decompression and fusion measures, i.e. considered the highest and lowest operated segment across both measures where applicable to estimate the total extent.

(e) BMI was recorded as a categorical factor in the 2011 version, while the 2017 version recorded it as an exact score based on patient's weight and height. We categorised the continuous BMI scores into <20, 20-25, 26-30, 31-35, and >35 to match the 2011 version data, as transformation in the opposite direction would not be possible. According to WHO classification, BMI <18.5 is considered underweight, <25 normal, >25 overweight, >30 obese, and >35 severely obese. From <20 category, it was not possible to distinguish between underweight and normal weight, therefore any BMI ≤25 was classified as normal. A previous study found no differences in spinal surgery outcomes between obese and extremely obese individuals with disc herniation or spinal stenosis [1], therefore, we combined BMI >30 into a single 'obese' category. Finally, BMI between 26 and 30 was classified as overweight.

Patients' age at the time of surgery was calculated by subtracting their year of birth (as full date of birth was not available, it was filled with the first day of the year) from intervention date.

Previous spinal surgeries included history of any spinal surgery on the same or different level.

Operation time was originally recorded as categories with 1-2 hours intervals (from <1 to 10), however, since there were few cases with duration beyond 4 hours, we collapsed the durations longer than 3 hours into a single category.

Similarly, blood loss was recorded as categories with 500-1000 ml intervals up to >2000 ml, but since cases with blood loss >500 ml were rather rare (1% of all cases), we collapsed them into a single category.

Duration of hospital stay was calculated by subtracting intervention date from discharge date. There were single values suggesting obvious data entry errors (e.g. durations > 1 year), as well as outliers, thus based on the distribution (i.e. its flattening with only single cases) and clinical judgement of what would constitute realistic length of stay, values of >84 days (3 months) duration were removed.

#### Methods S2: Handling missing data

##### Multivariate Imputation by Chained Equations (MICE)

In the MICE algorithm, each variable with missing data is modelled conditional on the other variables in the data (including outcome variables and any auxiliary variables that may be related to missingness – here e.g. the surgery form version later used to time-split the data into development and validation dataset was also included) using a series of regression models. The MICE algorithm allows to impute a mix of continuous and categorical variables, each according to its distribution, therefore predictive mean matching was used for numeric factors (in contrast to linear regression, it does not assume normal distribution and produces imputed values with a distribution more closely resembling that of observed values [2], logistic regression for categorical factors with two levels, and multinomial logit models for categorical factors with more than two levels. During the MICE procedure, missing values in each variable are first replaced with temporary placeholders based on available values for that variable. For one variable at a time, the placeholder values are set back to missing, and that variable is regressed on the remaining variables in the dataset. The fitted regression model is used to predict the missing values on the first variable. That imputed variable is then used as one of the independent variables in regression models for the remaining variables with missing data. In each cycle, all variables with any missing values are imputed with predictions that reflect any relationships observed between the variables in the data. This process undergoes multiple iterations (here 40) where the imputations of missing values on each variable are updated at every iteration, and eventually the final iteration is retained as the imputed dataset [3].

##### Multiple imputation diagnostics

Note that there was only 1 missing value on surgeon credentials, and 2-3 missing values on age and extent of surgery, therefore, it was not feasible to conduct all diagnostics on the imputation of these variables and some figures may not be shown.

The convergence of the algorithm was inspected by plotting trace lines of the mean and SD of the imputed values. Trace lines for all variables with missing values appeared to be free from any trends at later iterations (they oscillated within a narrow range of mean values), demonstrating good convergence (**Fig. S1**).

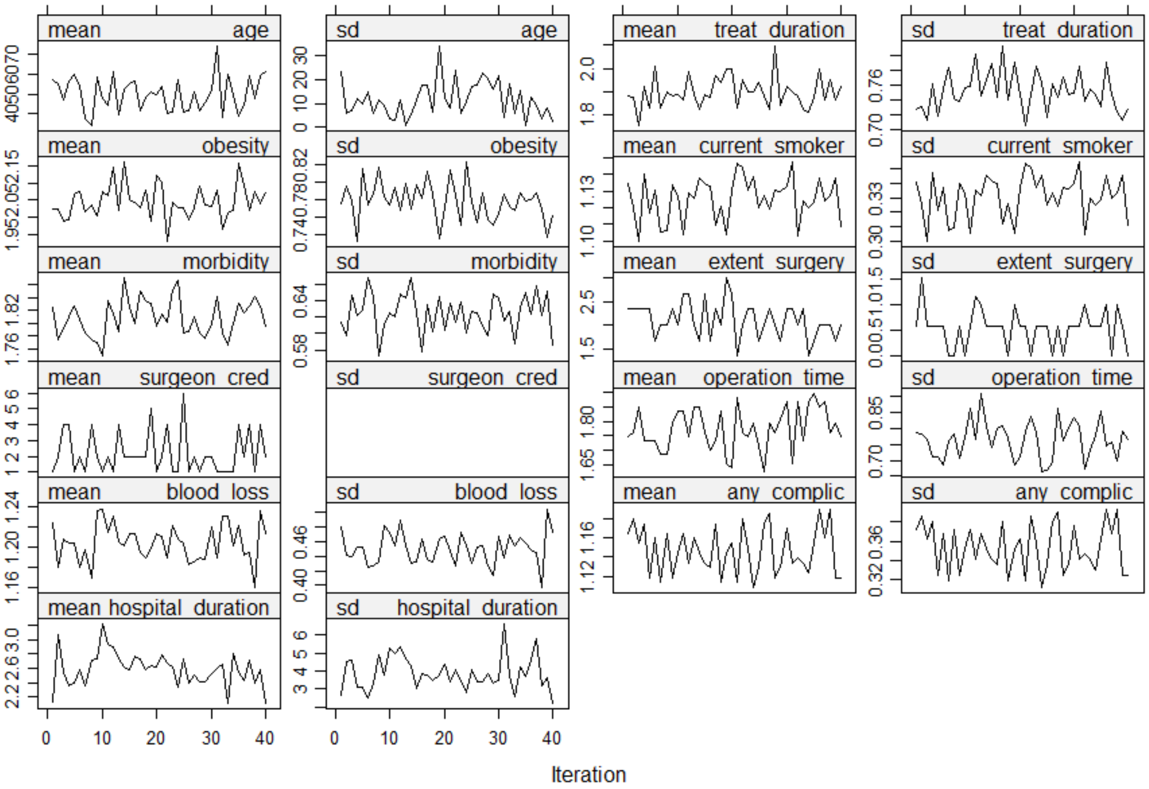

**Fig. S1.** Trace lines representing means and SDs of imputed values at each iteration.

As shown in **Fig. S2**, the imputed values were within the range of plausible values and their distributions closely corresponded to those of observed values.

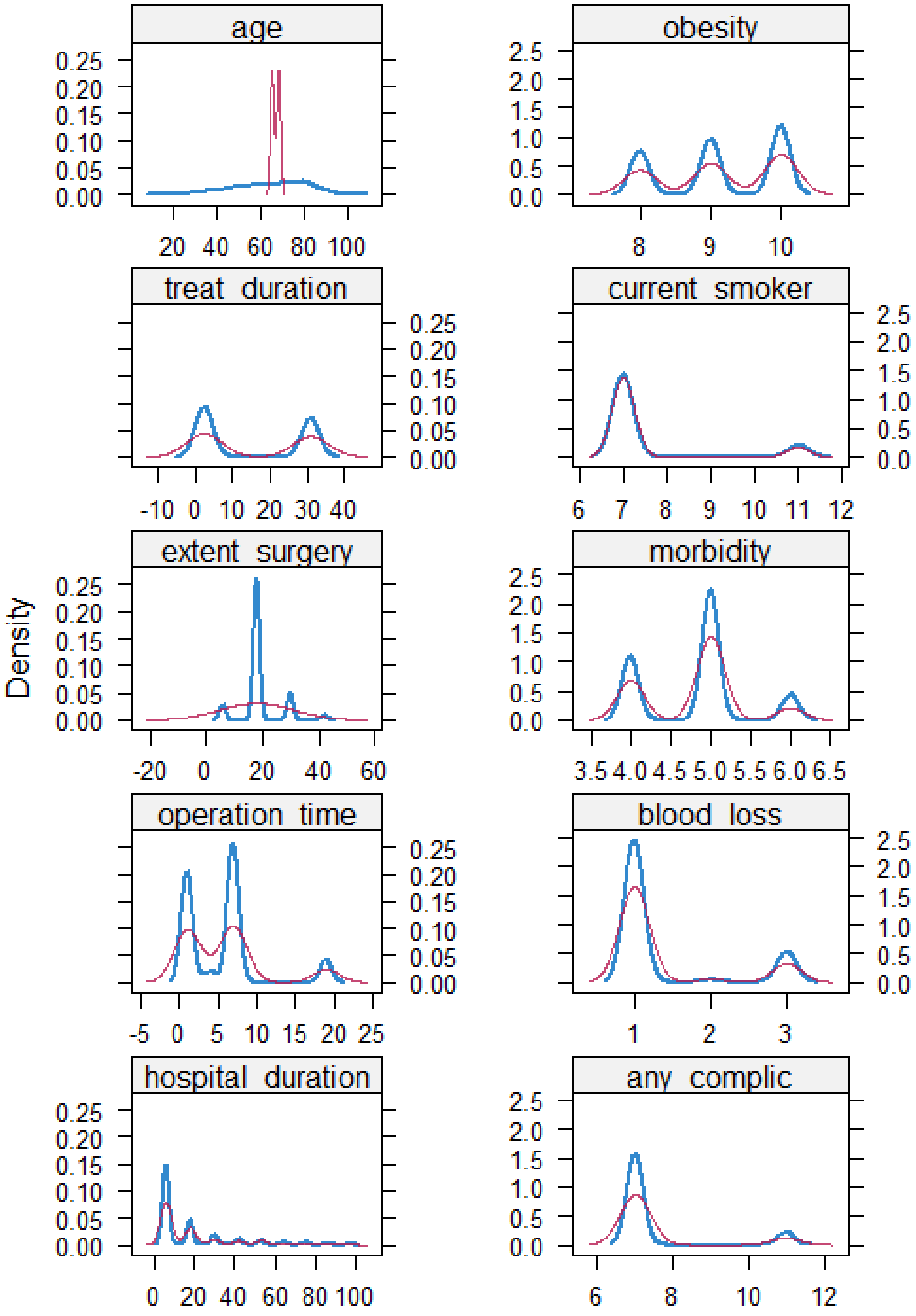

**Fig. S2.** Kernel density estimates for the marginal distributions of the observed data (blue) and the densities per variable calculated from the imputed data (red). Note that there were only 2-3 values were missing on ‘age’ and ‘extent of surgery’ resulting in different density distributions, yet still within expected values.

We further compared the distributions of observed and imputed data conditional on the missingness probability. That is, we regressed the probability of observations in each variable being missing on all variables in the imputed dataset, and then plotted each variable against its propensity score. Under the missing at random assumption, the conditional distributions should be similar if the assumed imputation models have good fits [4], as seen in **Fig. S3**.

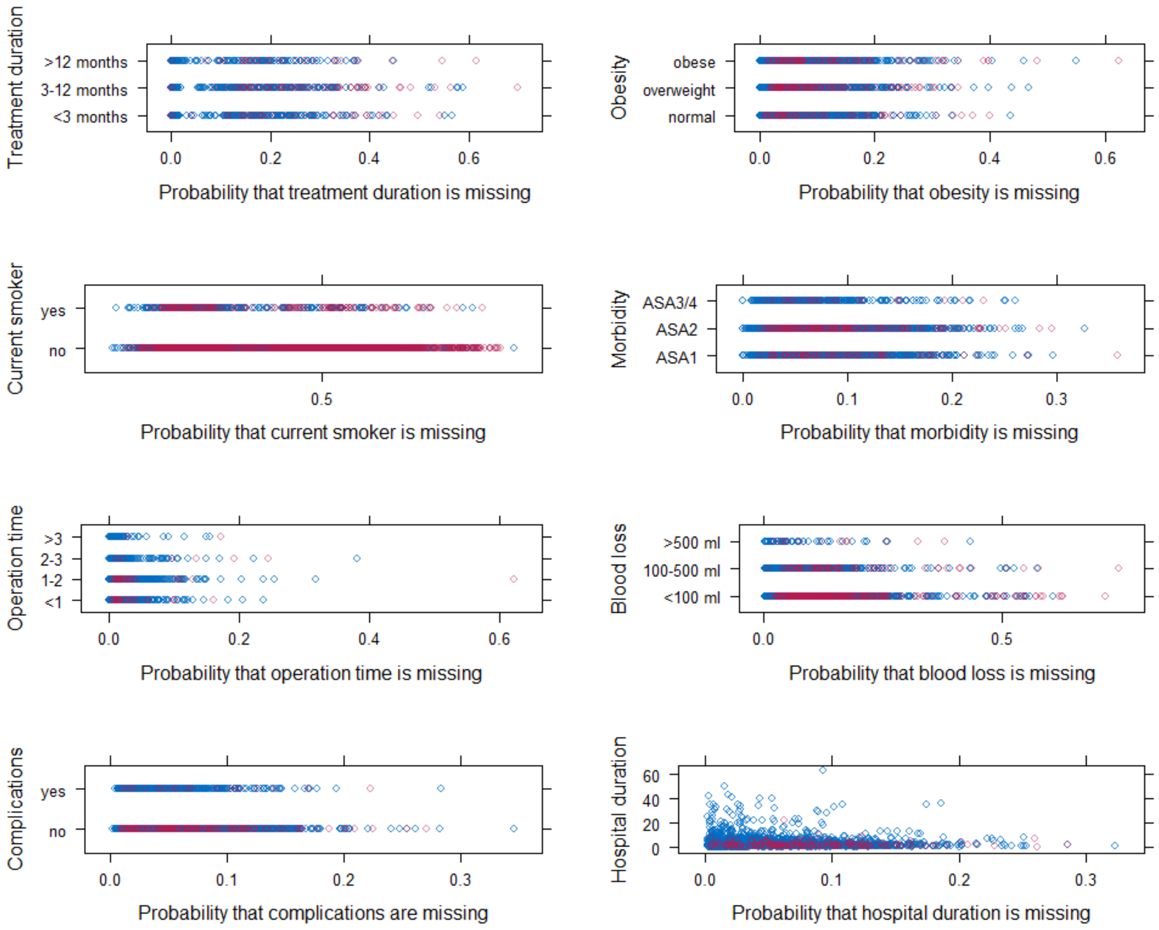

**Fig. S3.** Each variable with missing data plotted against its propensity score for observed (blue) and imputed (red) values.

We examined the residuals of the relationships between the imputed variables and their propensity scores. Under missing at random assumption we would expect the spread of the residuals to be similar for observed and imputed data, thus their distributions should overlap [4], as is apparent in **Fig. S4**.

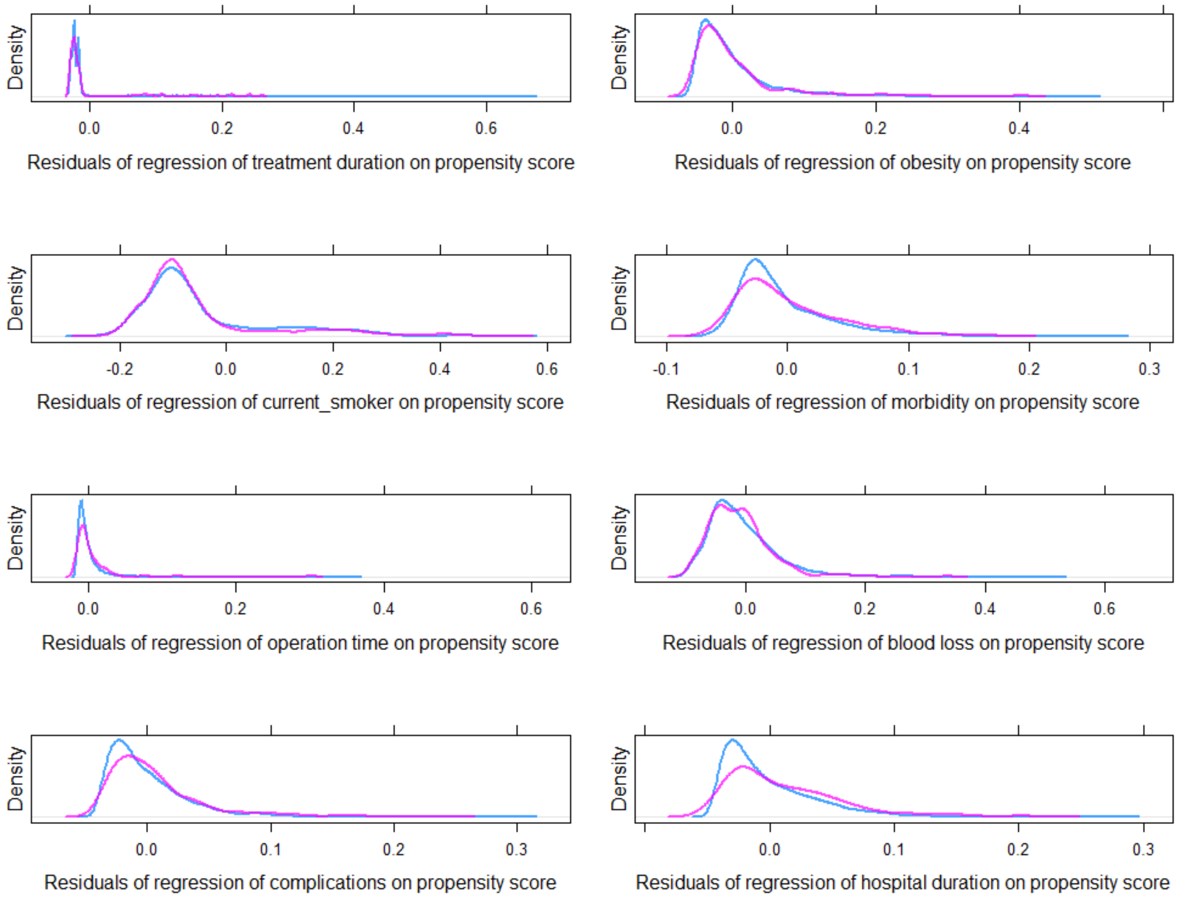

**Fig. S4.** Distributions of the residuals of regression models of each variable with missing data on its propensity score, grouped by observed (blue) and imputed (red) data.

To determine whether the data missingness could be predicted from any other variables present in the dataset, we fit logistic regression models on the probability of missingness for each variable with >3 missing values. Indeed, the probability of missing data on each factor was associated with several other factors included in the imputation models (**Table S1**), further supporting the assumption that data was missing at random.

**Table S1.** Results overview of the logistic regression models on the probability of missingness of each variable with missing data.

| *Predictors* | | *Treatment duration missing* | *BMI missing* | *Current smoker missing* | *Morbidity missing* | *Operation time missing* | *Blood loss missing* | *Complications missing* | *Hospital stay missing* |
|---|---|---|---|---|---|---|---|---|---|
| (Intercept) | | ● | ● | ● | ● | ● | ● | ● | ● |
| Age | | | ● | | | | ● | ● | ● |
| Gender (ref. female) | male | | | | | | | | |
| Degen. disease (ref. disc herniation) | stenosis | | ● | | | | ● | | |
| | disc herniation & stenosis | | ● | | | | ● | | |
| Previous surgeries (ref. no) | yes | ● | | | | | | | |
| Treatment duration (ref. <3 months) | 3-12 months | | ● | | ● | ● | | ● | ● |
| | >12 months | | ● | ● | ● | | | ● | ● |
| BMI (ref. normal) | overweight | | | | | | | | |
| | obese | | | | | | | | |
| Current smoker (ref. no) | yes | | | | | | | | |
| Morbidity (ref. ASA1) | ASA2 | | | | | | | | |
| | ASA3/4 | | | | | | ● | | |
| Baseline COMI | | | | | | | | | |
| Baseline back pain | | | | | | | ● | | |
| Baseline leg pain | | | ● | | | | | ● | |
| Extent of surgery (ref. 1) | 2 | | | ● | | | ● | | |
| | 3 | | | | | | | | |
| | 4+ | | | ● | | | | | |
| Surgical measures (ref. decompression) | decompression & fusion | | ● | ● | ● | ● | ● | | |
| Surgeon (ref. specialized spine) | board certified neuro | | ● | | | | ● | ● | ● |
| | board certified ortho | | | ● | | ● | ● | ● | ● |
| | neuro in training | | ● | | | | | ● | ● |
| | ortho in training | | | | | | | ● | ● |
| | other | | | ● | | ● | | | |
| Operation time (ref. <1) | 1-2 | | | ● | | | | | |
| | 2-3 | | ● | ● | | | | | |
| | >3 | | | ● | | | ● | | |

| | | | | | | | | | |
|---|---|---|---|---|---|---|---|---|---|
| Blood loss (ref. <100 ml) | 100-500 ml | | | | | ● | | | |
| | >500 ml | | | | | | | | |
| Complications (ref. no) | yes | | | | | | | | |
| (log)Hospital stay | | | ● | | | ● | | | |
| Follow-up (ref. <3 months) | 12 months | | | ● | ● | | | | |
| | 24 months | | | ● | ● | | | | |
| COMI change | | | | | | | | | |
| Back pain change | | ● | | | | | | | |
| Leg pain change | | | | | | | | | |
| COMI MCID (ref. no) | yes | | | | | | | | |
| Back pain MCID (ref. no) | yes | | | | | | | | |
| Leg pain MCID (ref. no) | yes | | | | | | | | |
| Surgery form version (ref. 2011) | 2017 | ● | | ● | ● | | ● | ● | ● |
| Nagelkerke $R^2$ | | 0.482 | 0.127 | 0.152 | 0.074 | 0.112 | 0.104 | 0.074 | 0.105 |
| AUC (95% CI) | | 0.95 (0.93, 0.97) | 0.74 (0.71, 0.77) | 0.69 (0.67, 0.71) | 0.69 (0.66, 0.72) | 0.76 (0.71, 0.81) | 0.69 (0.67, 0.72) | 0.70 (0.66, 0.74) | 0.74 (0.71, 0.78) |

*Note.* Each column corresponds to the outcome (probability of missing data on a particular variable) of each regression model, with the remaining variables in the dataset as potential predictors. Significant predictors are indicated by filled circles: ● p < .001, ● p < .01, ● p < .05.
95% CI, confidence interval; ASA, American Society of Anaesthesiologists; AUC, Area Under the Receiver-Operating Characteristic Curve; BMI, Body Mass Index; COMI, Core Outcome Measures Index.

#### Methods S3: Random forest hyperparameter tuning

Several hyperparameters of the RF algorithm can be tuned to maximise its predictive accuracy. The number of trees (*ntree*) determines how many decision trees should be constructed (and then aggregated) with the aim to stabilise the error rate (commonly it is set between 100 and 500 trees, or to 10 times the number of predictors). Increasing the number of trees provides more robust and stable error estimates and variable importance measures without increasing the risk of overfitting, therefore, we set each model to 500 trees. The number of predictors (p) to consider at each split (*mtry*) controls the split variable set randomisation. By default, it is calculated as p/3 for classification and √p for regression, however, a higher value may perform better when the data is noisy and there are fewer relevant predictors (on the contrary, a lower value might perform better when there are many relevant predictors). The minimum size of terminal nodes (*nodesize*) controls the depth or complexity of individual decision trees. It takes default values of 1 for classification and 5 for regression, however, performance in noisy data might improve with increased node size (i.e., decreased tree depth). Sample size (*sampsize*) determines the fraction of the observations selected for training (constructing) each tree and by default is set to 0.632. Decreasing this proportion reduces between-tree correlation, which can improve prediction accuracy. Replacement parameter (*replace*) controls whether bootstrap sampling of observations for training should be done with (default) or without replacement. While sampling without replacement would be more appropriate for evaluation of relative variable importance as it can minimise the bias towards continuous and categorical variables with higher number of levels, sampling with replacement is more appropriate for building predictive models (which is the primary aim of the present study).

Hyperparameter tuning was conducted on the development data using a *ranger* R package [5]. For each model, we performed a full Cartesian grid search with the possible hyperparameters presented in **Table S2**. The final hyperparameters for each outcome model were selected based on the lowest OOB error for classification and the lowest root mean square error (RMSE) for regression.

**Table S2.** Random forest hyperparameters.

| ***Hyper-parameters*** | ***Tested values*** | ***Final values*** | | | | | |
|---|---|---|---|---|---|---|---|
| | | *COMI MCID* | *Back pain MCID* | *Leg pain MCID* | *COMI change* | *Back pain change* | *Leg pain change* |
| *ntrees* | 500 | 500 | 500 | 500 | 500 | 500 | 500 |
| *mtry* | 3, 4, **6**, 9, 12 (classification) / 1, 3, **4**, 6, 8 (regression) | 6 | 4 | 4 | 12 | 8 | 8 |
| *min.node.size (nodesize)* | **1**, 3, **5**, 10, 20 | 3 | 1 | 5 | 20 | 20 | 20 |
| *replace* | **TRUE**, FALSE | TRUE | TRUE | FALSE | TRUE | TRUE | TRUE |
| *sample.fraction (sampsize)* | 0.5, **0.63**, 0.8 | 0.63 | 0.5 | 0.5 | 0.5 | 0.5 | 0.5 |

*Note*. In brackets, hyperparameter names in *randomForest* corresponding to those in *ranger* function. In bold, default values in *randomForest* function.

*ntree*, number of trees; *mtry,* number of predictors considered at each split; *nodesize,* minimum size of terminal nodes; *replace*, sampling training observations with vs. without replacement; *sampsize*, proportion of all observations sampled for training; COMI, Core Outcome Measures Index; MCID, Minimal Clinically Important Difference.

#### Results S1: Participants

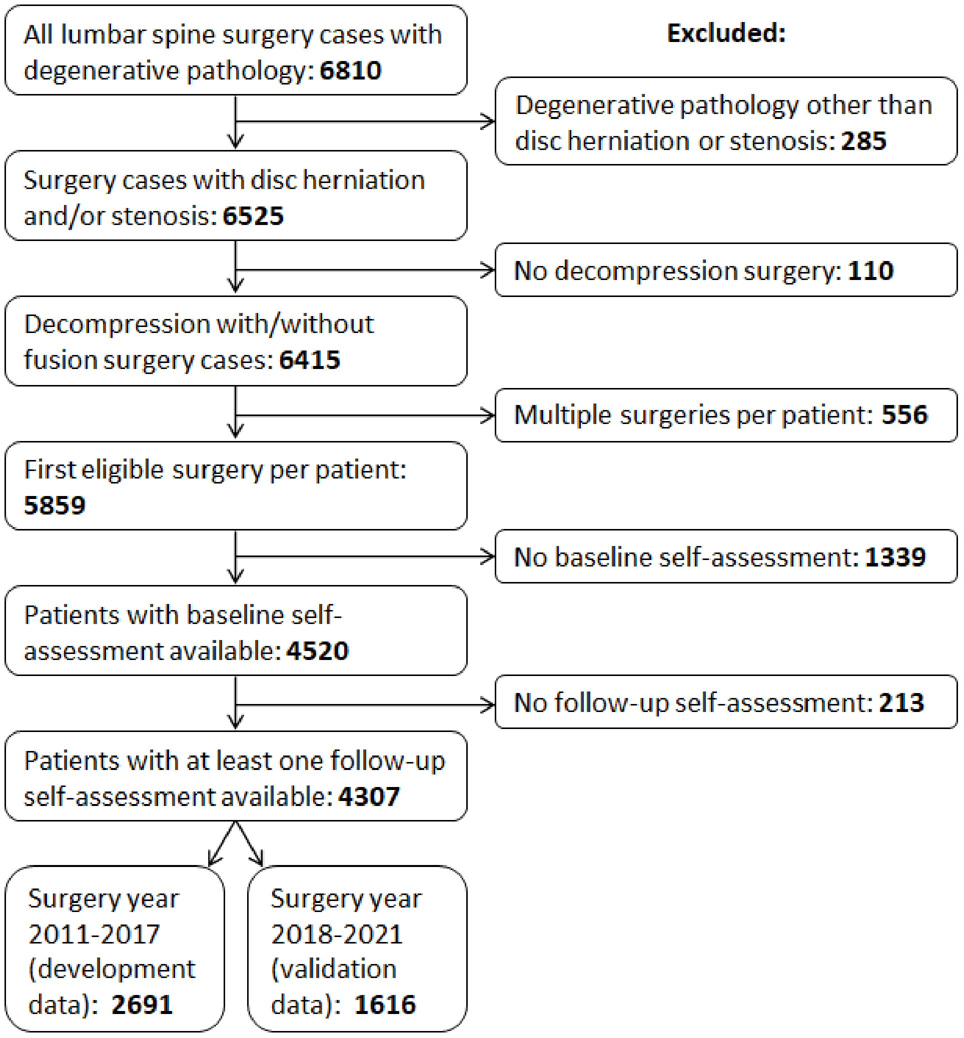

**Fig. S5.** The flow of surgery cases and participants through the selection process.

##### Included versus excluded sample characteristics

**Table S3** presents descriptive characteristics of eligible participants included in the analysis and those who were excluded due to incomplete baseline and/or follow-up self-assessments (but who would be otherwise eligible), with results of between-group comparisons on candidate predictor variables (*t*-tests for continuous, and $\chi^2$ tests for categorical factors). Excluded participants were on average 10 years younger, stayed in the hospital for 0.8 days less, but did not differ from included participants on the baseline COMI, back pain, and leg intensity (where data were available). Among the excluded participants, disc herniation was more, and stenosis less common relative to the included sample. Excluded participants tended to have shorter duration of previous treatment, and smaller proportion on non-smokers (but greater proportion of missing values on that factor). There was also greater proportion of participants with no morbidity and smaller proportion of those with mild/moderate morbidity compared to included participants. A greater proportion of excluded participants underwent surgery spanning 2 levels, and a smaller proportion had 3 or more levels operated. Operation time in excluded participants tended to be longer, with a smaller proportion of surgeries lasting <1 hour. Decompression alone was a more, and decompression with fusion a less common type of surgery among the excluded participants. Excluded participants were more often operated by board-certified neurosurgeons and neurosurgeons in training, and less often by specialised spine surgeons, compared to the included participants. There were no significant differences on other predictor variables and proportions of missing predictor data were comparable between included and excluded participants, except for the smoking status and BMI, with more missingness in the excluded patient sample. Despite some statistically significant differences, the complete sample data covers the full range of possible predictor values observed in the incomplete sample and contains sufficient number of observations across the level of categorical factors to conclude that the subgroups of the population of interest are well represented in the included data.

Within the included patient sample, among those with spinal stenosis, 68% had lateral, 60% central, and 30% foraminal stenosis (note that the percentages do not add up to 100% as often more than one option was recorded for the same patient). Pain relief was the most common surgical goal (97%; 16% axial, 95% peripheral), followed by functional (30%), motor (11%), and sensory (8%) improvement. The most common decompression measures included flavectomy (54%), laminectomy (43%), discectomy (41%), foraminotomy (39%), laminotomy (32%), and facet joint resection (27%). Fusion surgeries most often involved transforaminal interbody (51%), posterolateral (48%), posterior (18%), and posterior interbody fusion (14%). Out of those patients with a history of previous spinal surgery, 75% underwent one, 19% two, and 6% three or more previous surgeries.

**Table S3.** Characteristics of included and excluded participants on continuous and categorical factors.

| | | *Complete (N = 4307)* | | | *Incomplete (N = 1552)* | | | *Difference* | | |
|---|---|---|---|---|---|---|---|---|---|---|
| ***Factor*** | | *Mean (SD)* | *Range* | *NA (%)* | *Mean (SD)* | *Range* | *NA (%)* | *t* | *df* | *p* |
| Age (years) | | 59.70 (14.91) | 18 - 94 | 2 (<1%) | 49.80 (15.92) | 17 - 89 | 4 (<1%) | -21.32 | 2585 | <0.001 |
| Hospital duration (days) | | 2.52 (3.92) | 0 - 63 | 192 (4%) | 3.34 (6.39) | 0 - 80 | 109 (7%) | 4.63 | 1837 | <0.001 |
| Baseline COMI (0-10) | | 8.14 (1.61) | 0 - 10 | 0 | 8.25 (1.55) | 1 - 10 | 1339 (86%) | 1.04 | 235 | 0.301 |
| Baseline back pain (0-10) | | 6.67(2.73) | 0 - 10 | 0 | 6.79 (2.74) | 0 - 10 | 1339 (86%) | 0.63 | 233 | 0.526 |
| Baseline leg pain (0-10) | | 7.82 (2.19) | 0 - 10 | 0 | 7.74 (2.45) | 0 - 10 | 1339 (86%) | -0.47 | 229 | 0.640 |
| COMI change (0-10) | | 3.27 (3.11) | -6 - 10 | 0 | | | | | | |
| Back pain change (0-10) | | 2.28 (3.38) | -10 -10 | 0 | | | | | | |
| Leg pain change (0-10) | | 3.78 (3.71) | -9 - 10 | 0 | | | | | | |
| ***Factor*** | ***Levels*** | *N* | *%* | | *N* | *%* | | $\chi^2$ | *df* | *p* |
| Degenerative disease | disc herniation | 1041 | 24% | | 670 | 43% | | 342.41 | 2 | <0.001 |
| | stenosis | 2413 | 56% | | 452 | 29% | | | | |
| | disc herniation & stenosis | 853 | 20% | | 430 | 28% | | | | |
| Previous surgeries | no | 3478 | 81% | | 1247 | 80% | | 0.09 | 1 | 0.758 |
| | yes | 829 | 19% | | 305 | 20% | | | | |
| Duration of treatment | <3 months | 1159 | 27% | | 683 | 44% | | 177.81 | 2 | <0.001 |
| | 3-12 months | 1801 | 42% | | 549 | 35% | | | | |
| | >12 months | 1228 | 29% | | 264 | 17% | | | | |
| | NA | 119 | 3% | | 56 | 4% | | | | |
| BMI | normal | 1047 | 24% | | 369 | 24% | | 1.29 | 2 | 0.525 |
| | overweight | 1652 | 38% | | 536 | 35% | | | | |
| | obese | 1358 | 32% | | 443 | 29% | | | | |
| | NA | 250 | 6% | | 204 | 13% | | | | |
| Current smoker | no | 2430 | 56% | | 717 | 46% | | 8.68 | 1 | 0.003 |
| | yes | 348 | 8% | | 142 | 9% | | | | |
| | NA | 1529 | 36% | | 693 | 45% | | | | |
| Morbidity | ASA1 | 1171 | 27% | | 613 | 39% | | 88.06 | 2 | <0.001 |
| | ASA2 | 2387 | 55% | | 693 | 45% | | | | |
| | ASA3/4 | 471 | 11% | | 134 | 9% | | | | |
| | NA | 278 | 6% | | 112 | 7% | | | | |

| | | | | | | | | |
|---|---|---|---|---|---|---|---|---|
| Gender | female | 2136 | 50% | 753 | 49% | 0.46 | 1 | 0.499 |
| | male | 2171 | 50% | 798 | 51% | | | |
| | NA | 0 | 0% | 1 | 0% | | | |
| Extent of surgery | 1 | 340 | 8% | 130 | 8% | 48.81 | 3 | <0.001 |
| | 2 | 3231 | 75% | 1265 | 82% | | | |
| | 3 | 622 | 14% | 142 | 9% | | | |
| | ≥4 | 111 | 3% | 12 | 1% | | | |
| | NA | 3 | 0% | 3 | 0% | | | |
| Operation time | <1 | 1648 | 38% | 518 | 33% | 28.07 | 3 | <0.001 |
| | 1-2 | 2077 | 48% | 773 | 50% | | | |
| | 2-3 | 341 | 8% | 176 | 11% | | | |
| | >3 | 174 | 4% | 43 | 3% | | | |
| | NA | 67 | 2% | 42 | 3% | | | |
| Blood loss | <100 ml | 3142 | 73% | 1121 | 72% | 1.31 | 2 | 0.520 |
| | 100-500 ml | 677 | 16% | 229 | 15% | | | |
| | >500 ml | 56 | 1% | 15 | 1% | | | |
| | NA | 432 | 10% | 187 | 12% | | | |
| Complications | no | 3610 | 84% | 1256 | 81% | 0.02 | 1 | 0.892 |
| | yes | 503 | 12% | 178 | 11% | | | |
| | NA | 194 | 5% | 118 | 8% | | | |
| Surgical measures | decompression | 4011 | 93% | 1492 | 96% | 17.55 | 1 | <0.001 |
| | decompression & fusion | 296 | 7% | 60 | 4% | | | |
| Surgeon credentials | specialized spine | 1409 | 33% | 345 | 22% | 73.47 | 5 | <0.001 |
| | board certified neuro | 1875 | 44% | 736 | 47% | | | |
| | board certified ortho | 101 | 2% | 36 | 2% | | | |
| | neuro in training | 818 | 19% | 402 | 26% | | | |
| | ortho in training | 27 | 1% | 6 | 0% | | | |
| | other | 76 | 2% | 24 | 2% | | | |
| | NA | 1 | 0% | 3 | 0% | | | |
| Follow-up interval | 3 months | 368 | 9% | | | | | |
| | 12 months | 901 | 21% | | | | | |
| | 24 months | 3038 | 71% | | | | | |
| COMI MCID | FALSE | 1829 | 42% | | | | | |

| | | | |
|---|---|---|---|
| | TRUE | 2478 | 58% |
| Back pain MCID | FALSE | 1976 | 46% |
| | TRUE | 2331 | 54% |
| Leg pain MCID | FALSE | 1420 | 33% |
| | TRUE | 2887 | 67% |

*Note*. Continuous outcomes, MCID outcomes, and follow-up interval are presented only for complete sample. Baseline COMI, back, and leg pain scores are presented for the portion of incomplete sample that had available data from the preoperative self-assessment (N = 213).
ASA, American Society of Anaesthesiologists morbidity class; BMI, body mass index; COMI, Core Outcome Measures Index; MCID, Minimal Clinically Important Difference; NA, number of missing values; neuro, neurosurgeon; ortho, orthopaedic surgeon.

The magnitude of change in COMI, back, or leg pain, or the proportion of patients achieving MCID on these measures in the included sample, did not depend on the follow-up interval (**Fig. S6**). This factor was nonetheless included as a potential confounder in the prediction models, however, it did not significantly predict any of the outcomes.

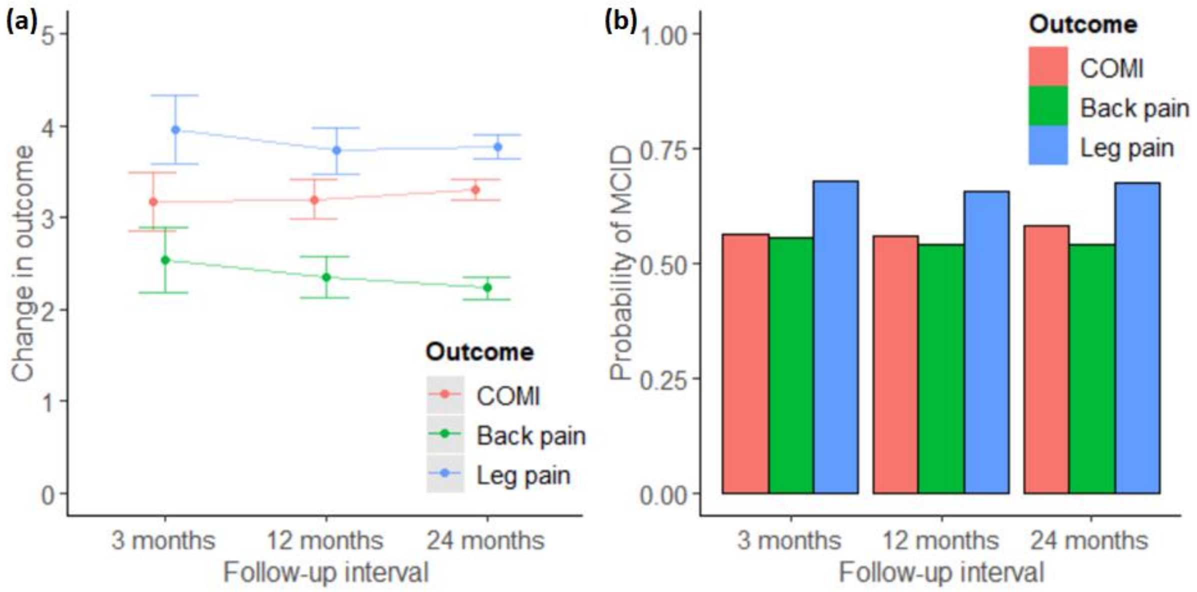

**Fig. S6.** (a) Mean with 95% CI change in COMI, back pain, and leg pain outcomes at each follow-up interval. (b) Proportions of patients who achieved MCID on these outcomes at each follow-up interval. 3 months N = 368, 12 months N = 901, 24 months N = 3038.

#### Development versus validation sample characteristics

Patient characteristics and group differences between the development and validation samples are presented in **Table S4**. Participants in the development sample were on average 3 years older, had 0.1 lower baseline COMI score, 0.2 lower back pain intensity, achieved 0.4 lower reduction in back pain, and 0.4 lower reduction in leg pain than participants in the validation sample. Compared to the validation sample, the development sample was also characterised by a greater proportion of patients with spinal stenosis alone and a smaller proportion of those with disc herniation and stenosis; a greater proportion of patients with shorter (<3 and 3-12 months) duration of previous treatment and a smaller proportion of those with >12 months duration; a greater proportion of overweight and a smaller proportion of obese individuals; a greater proportion of smokers; a greater proportion of patients with severe morbidity and a smaller proportion of those without morbidity; a greater proportion of males; more surgeries lasting <1 hour and less lasting 2 or more hours; a greater proportion of patients with <100 ml blood loss and a smaller proportion of those with >500 ml blood loss; a greater proportion of complications; more surgeries performed by neurosurgeons in training, orthopaedic surgeons in training, or other, and less surgeries performed by board certified neurosurgeons or orthopaedic surgeons; and a greater proportion of patients with the last follow-up at 24 months interval and a smaller proportion of shorter follow-up intervals. There were no differences between the development and validation samples in the proportion of patients with a history of previous surgery or those achieving MCID on any of the outcomes.

While our prospective minimum sample size estimation was based on the existing rules-of-thumb taking into account the number of candidate predictor parameters, subsequently published methods suggested additionally considering the total number of participants, outcome rates or mean outcome values in the population of interest, and the expected predictive performance of the models. Therefore, we conducted post-hoc calculations, using *pmsampsize* R package [6], to evaluate whether the available sample size would be considered sufficient based on the observed outcomes and models performance. The estimated minimum model development samples were 2887 for COMI MCID, 1625 for back pain MCID, 1625 for leg pain MCID, 1866 for COMI change, 1093 for back pain change, and 1109 for leg pain change scores. Therefore, our development dataset of 2691 patients exceeded the minimum required sample size for all outcomes except COMI MCID, for which it nearly reached the post-hoc estimate. Our external validation dataset of 1616 patients with 916-1092 outcome events also exceeded the minimum suggested number of events in validation samples for dichotomous outcomes (100; [7]), and based on post-hoc calculations, minimum sample size required to precisely estimate R2 values (684-890), calibration slope (1199 for back pain and 1212 for leg pain; except for COMI change, which would require 2165 patients), or residual variances (235) in external validation of prediction models for continuous outcomes [8].

**Table S4.** Participant characteristics and differences between the development and validation samples.

| | | ***Development data (N = 2691)*** | | ***Validation data (N = 1616)*** | | ***Difference*** | | |
|---|---|---|---|---|---|---|---|---|
| ***Continuous factors*** | | *Mean (SD)* | *Range* | *Mean (SD)* | *Range* | *t* | *df* | *p* |
| Age | | 60.95 (14.50) | 18- 94 | 57.62 (15.34) | 20 - 88 | 7.04 | 3252 | <0.001 |
| Baseline COMI | | 8.09 (1.55) | 0 - 10 | 8.21 (1.70) | 0 - 10 | -2.19 | 3158 | 0.028 |
| Baseline back pain | | 6.58 (2.68) | 0 - 10 | 6.80 (2.81) | 0 - 10 | -2.53 | 3266 | 0.012 |
| Baseline leg pain | | 7.78 (2.13) | 0 - 10 | 7.88 (2.27) | 0 - 10 | -1.38 | 3233 | 0.167 |
| Hospital duration | | 2.55 (3.92) | 0 - 63 | 2.41 (3.77) | 0 - 43 | 1.17 | 3505 | 0.240 |
| COMI change | | 3.22 (3.07) | -6.35 - 10 | 3.34 (3.18) | -6.1 - 10 | -1.15 | 3305 | 0.248 |
| Back pain change | | 2.15 (3.30) | -10 - 10 | 2.51 (3.50) | -8 - 10 | -3.30 | 3248 | 0.001 |
| Leg pain change | | 3.68 (3.69) | -9 - 10 | 3.94 (3.75) | -9 - 10 | -2.18 | 3355 | 0.029 |
| ***Categorical factors*** | ***Levels*** | *N* | *%* | *N* | *%* | $\chi^2$ | *df* | *p* |
| Degenerative disease | disc herniation | 621 | 23% | 420 | 26% | 27.95 | 2 | <0.001 |
| | stenosis | 1588 | 59% | 825 | 51% | | | |
| | disc herniation & stenosis | 482 | 18% | 371 | 23% | | | |
| Previous surgeries | no | 2166 | 80% | 1312 | 81% | 0.27 | 1 | 0.601 |
| | yes | 525 | 20% | 304 | 19% | | | |
| Duration of treatment | <3 months | 945 | 35% | 250 | 15% | 387.51 | 2 | <0.001 |
| | 3-12 months | 1226 | 46% | 631 | 39% | | | |
| | >12 months | 520 | 19% | 735 | 45% | | | |
| BMI | normal | 717 | 27% | 394 | 24% | 34.18 | 2 | <0.001 |
| | overweight | 1161 | 43% | 595 | 37% | | | |
| | obese | 813 | 30% | 627 | 39% | | | |
| Current smoker | no | 2314 | 86% | 1479 | 92% | 28.88 | 1 | <0.001 |
| | yes | 377 | 14% | 137 | 8% | | | |
| Morbidity | ASA1 | 736 | 27% | 517 | 32% | 26.56 | 2 | <0.001 |
| | ASA2 | 1598 | 59% | 960 | 59% | | | |
| | ASA3/4 | 357 | 13% | 139 | 9% | | | |
| Gender | female | 1297 | 48% | 839 | 52% | 5.44 | 1 | 0.020 |
| | male | 1394 | 52% | 777 | 48% | | | |
| Extent of surgery | 1 | 72 | 3% | 268 | 17% | 269.74 | 3 | <0.001 |
| | 2 | 2147 | 80% | 1087 | 67% | | | |
| | 3 | 402 | 15% | 220 | 14% | | | |

| | | | | | | | | |
|---|---|---|---|---|---|---|---|---|
| | ≥4 | 70 | 3% | 41 | 3% | | | |
| Operation time | <1 | 1130 | 42% | 546 | 34% | 61.52 | 3 | <0.001 |
| | 1-2 | 1306 | 49% | 801 | 50% | | | |
| | 2-3 | 175 | 7% | 173 | 11% | | | |
| | >3 | 80 | 3% | 96 | 6% | | | |
| Blood loss | <100 ml | 2213 | 82% | 1281 | 79% | 15.63 | 2 | <0.001 |
| | 100-500 ml | 450 | 17% | 295 | 18% | | | |
| | >500 ml | 28 | 1% | 40 | 2% | | | |
| Complications | no | 2300 | 85% | 1481 | 92% | 35.35 | 1 | <0.001 |
| | yes | 391 | 15% | 135 | 8% | | | |
| Surgical measures | decompression | 2545 | 95% | 1466 | 91% | 22.87 | 1 | <0.001 |
| | decompression & fusion | 146 | 5% | 150 | 9% | | | |
| Surgeon credentials | specialized spine | 890 | 33% | 519 | 32% | 90.60 | 4 | <0.001 |
| | board certified neuro | 1102 | 41% | 774 | 48% | | | |
| | board certified ortho | 28 | 1% | 73 | 5% | | | |
| | neuro in training | 570 | 21% | 248 | 15% | | | |
| | ortho in training | 25 | 1% | 2 | 0% | | | |
| | other | 76 | 3% | 0 | 0% | | | |
| Follow-up interval | 3 months | 104 | 4% | 264 | 16% | 828.58 | 2 | <0.001 |
| | 12 months | 272 | 10% | 629 | 39% | | | |
| | 24 months | 2315 | 86% | 723 | 45% | | | |
| COMI MCID | FALSE | 1144 | 43% | 685 | 42% | <0.01 | 1 | 0.962 |
| | TRUE | 1547 | 57% | 931 | 58% | | | |
| Back pain MCID | FALSE | 1276 | 47% | 700 | 43% | 2.09 | 1 | 0.148 |
| | TRUE | 1415 | 53% | 916 | 57% | | | |
| Leg pain MCID | FALSE | 896 | 33% | 524 | 32% | 0.15 | 1 | 0.703 |
| | TRUE | 1795 | 67% | 1092 | 68% | | | |

ASA, American Society of Anaesthesiologists morbidity class; BMI, body mass index; COMI, Core Outcome Measures Index; MCID, Minimal Clinically Important Difference; neuro, neurosurgeon; ortho, orthopaedic surgeon.

#### Results S2: Regression diagnostics

Pearson correlation coefficients between continuous candidate predictors in the development data did not exceed 0.7. Relationships between continuous predictors and continuous outcomes did not appear to deviate from linearity (**Fig. S7**). For logistic regression models, the relationships between the continuous predictor and logit of outcomes in the development data did not show severe deviations from linearity, although there were potential non-linear relationships between COMI MCID and baseline back pain and logarithm of the duration of hospital stay, and between leg pain MCID and baseline back pain (**Fig. S8**).

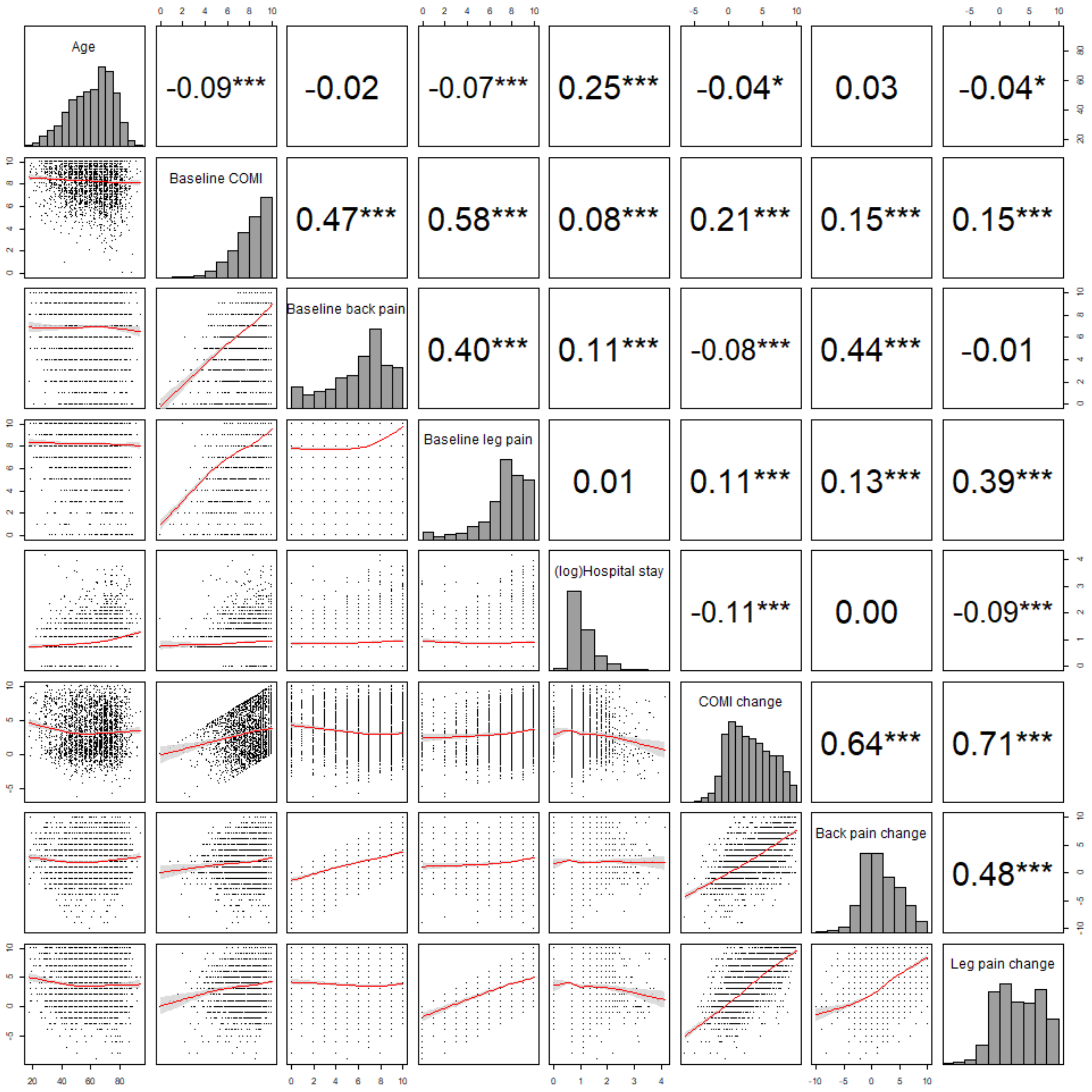

**Fig. S7.** Scatterplots with fitted smooth loess regression line (red) with 95% confidence interval (grey) (left-lower panel) and Pearson's correlation coefficients (right-upper panel) illustrating any relationships among continuous predictors (upper 5 rows) and between continuous predictors and outcomes (bottom 3 rows). Histograms illustrate distributions of the continuous variables. N = 2691. ***p<.001, **p<.01.

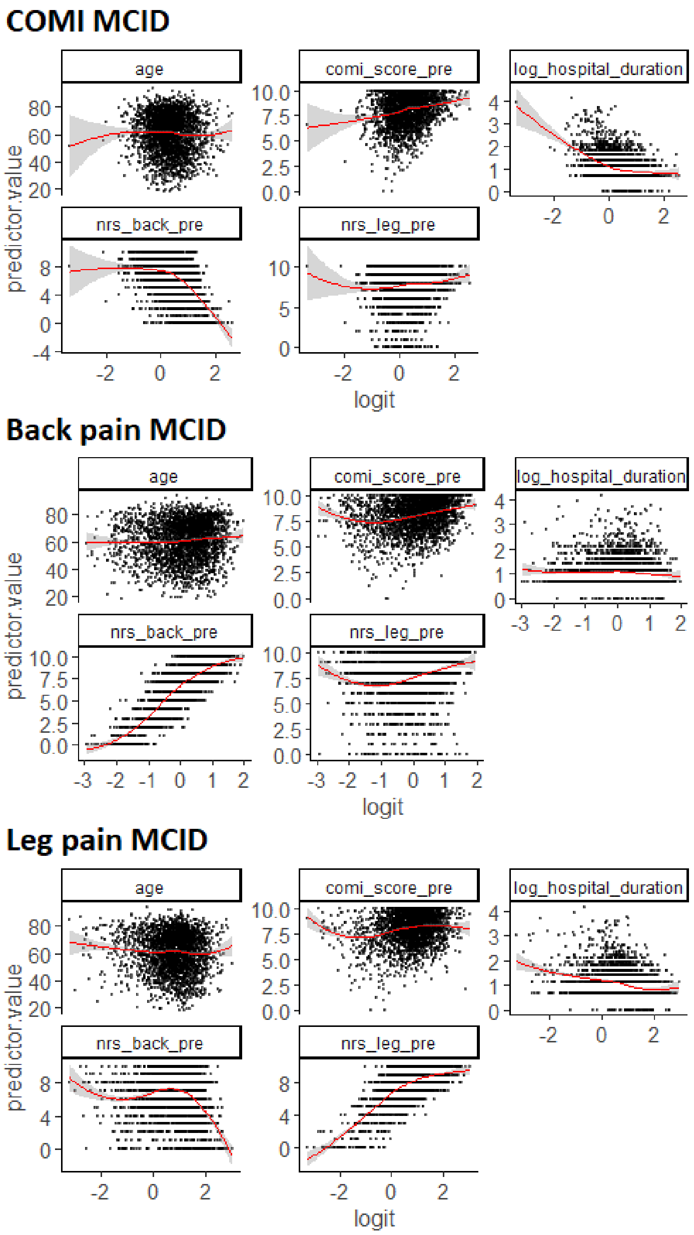

**Fig. S8.** Scatterplots with fitted smooth loess regression line (red) with 95% confidence interval (grey) illustrating the relationships between each continuous predictor and logit of outcome in the development data (N = 2691). comi_score_pre, baseline COMI; nrs_back_pre, baseline back pain; nrs_leg_pre, baseline leg pain.

#### Logistic regression

Variance inflation factors across all predictors for each outcome were <2.5, indicating there was no multicollinearity (values >5 would be problematic). As shown in **Fig. S9**, deviance residuals plotted against observation number did not exceed an absolute value of 3 and did not suggest a poor fit for any particular observations. Deviance residuals plotted against predicted probabilities showed patterns typical of binomial outcome distributions and suggested adequate model fits without highly influential outliers. Cook's distance values were also consistently <1 (COMI <0.007, backpain <0.005, leg pain <0.008) suggesting no highly influential cases.

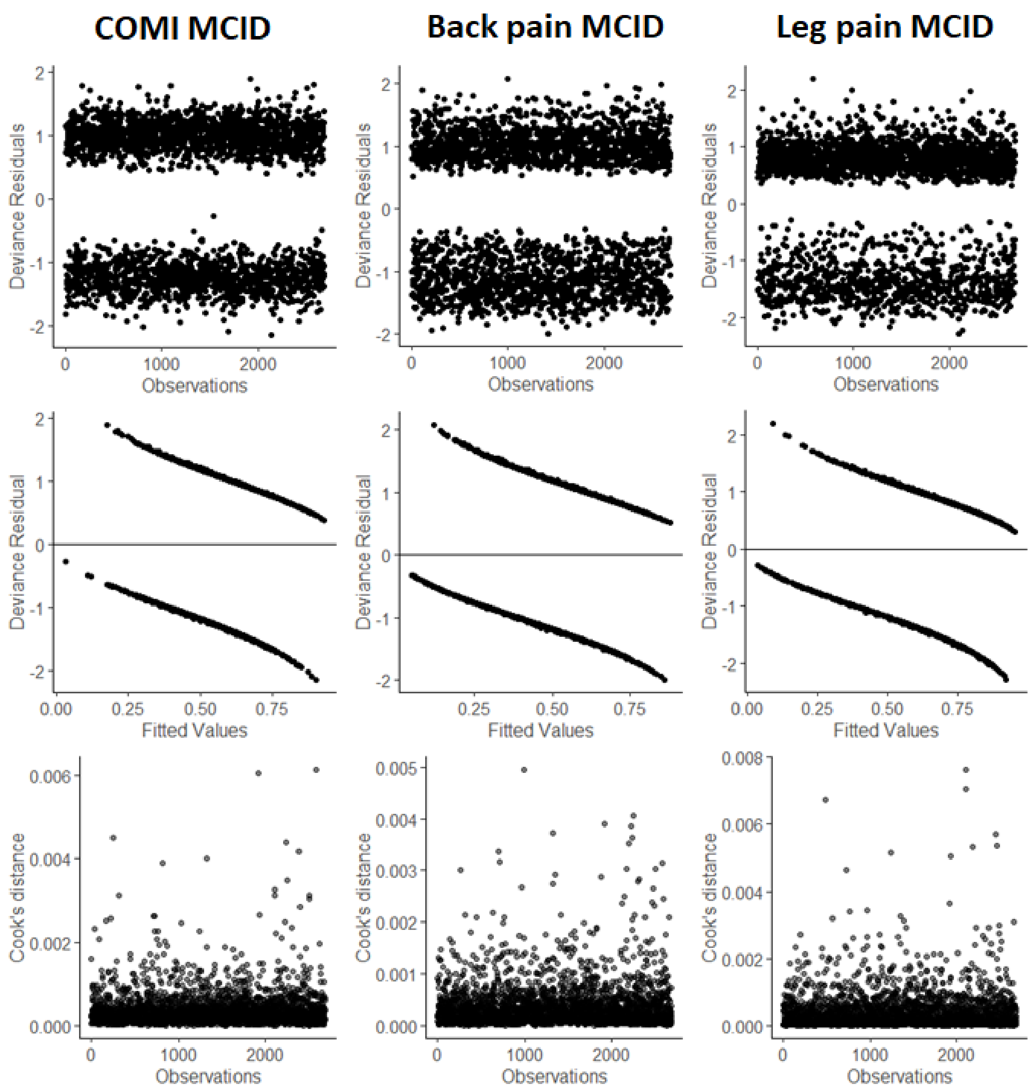

**Fig. S9.** Diagnostic plots for logistic regression models on Minimal Clinically Important Difference (MCID) in Core Outcome Measures Index (COMI), back pain, and leg pain. Deviance residuals are plotted against observation number (top panel), predicted outcome probabilities (middle panel), and Cook's distance is plotted against observation number (bottom panel).

#### Linear regression

As illustrated in the component residual plots (**Fig. S10**), the predictors appeared to have linear relationships with each outcome. For continuous predictors, there was substantial overlap between the least-squares residual lines and the best loess fit component lines. Boxplots for categorical predictors further illustrate homogeneity of variance across different levels of each factor.

(a) COMI change

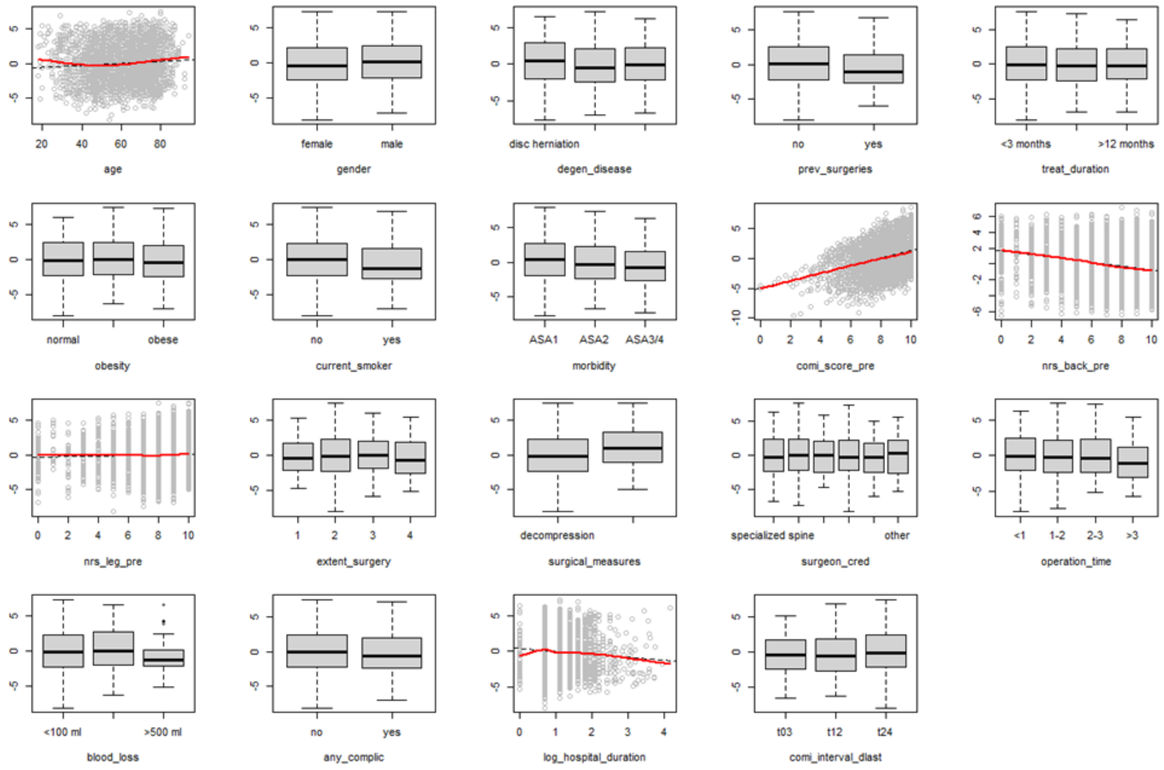

(b) Back pain change

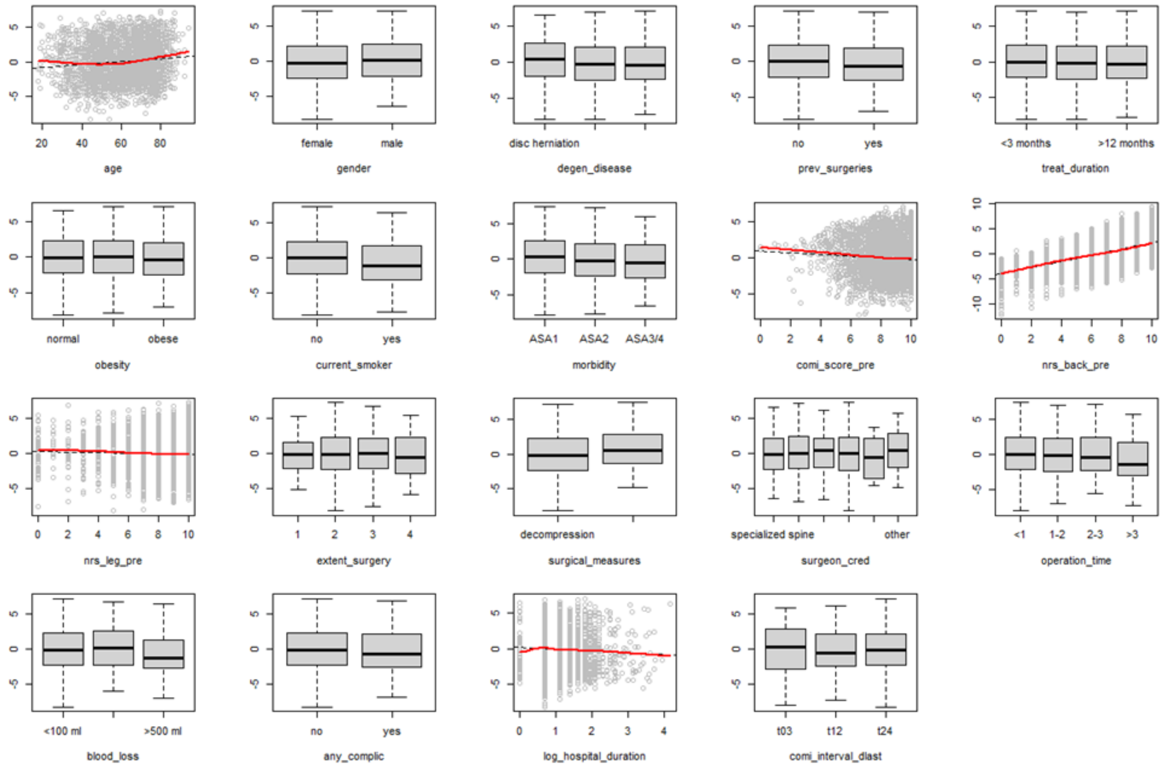

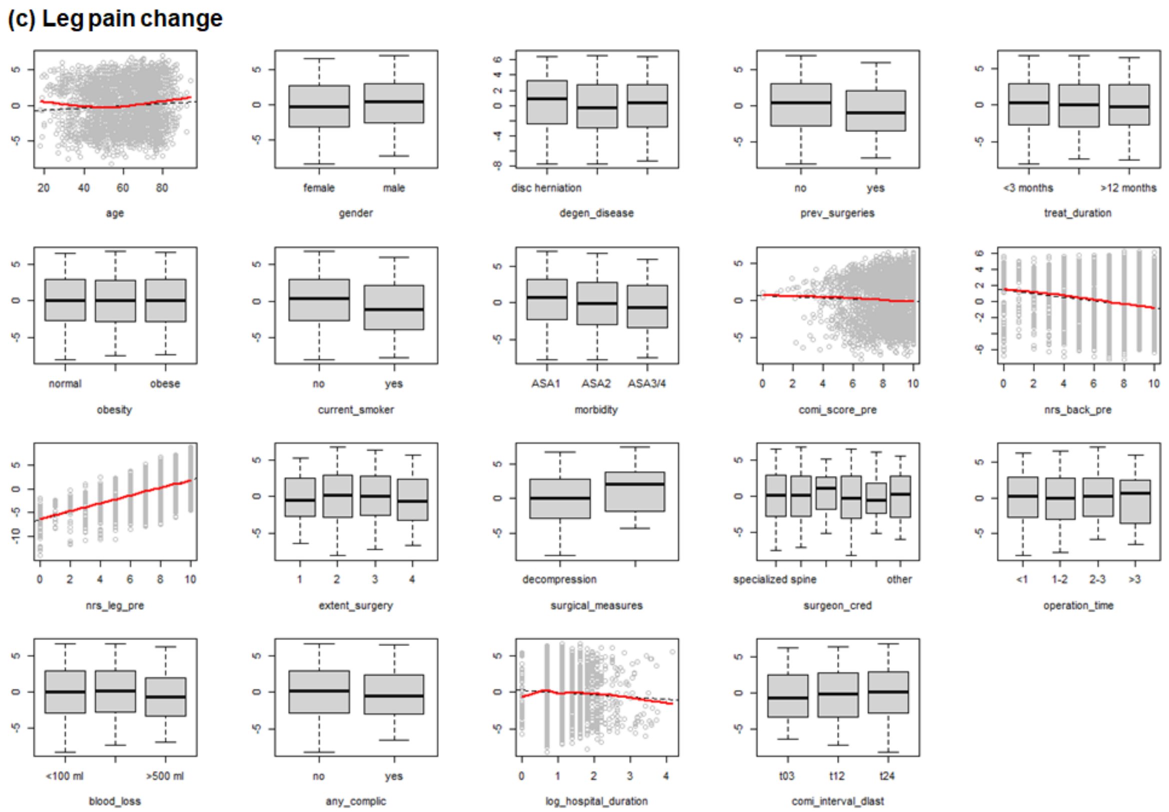

**Fig. S10.** Component residual plots. For continuous predictors, the black dotted least-squares lines model the residuals of each predictor against the outcome, and the red component lines represent the best loess fits. Substantial overlap between the residual line and the component line indicates a linear relationship between each predictor and the outcome variable: (a) change in Core Outcome Measures Index (COMI), (b) back pain, and (c) leg pain. Boxplots for categorical predictors illustrate residual variance across different levels of each factor.

There was no multicollinearity among the predictors, as variance inflation factors were ≤2.44. Standardised residuals did not exceed an absolute value of 3 (max. abs. COMI 2.84, back pain 2.82, leg pain 2.51), and when plotted against the predicted outcome values (**Fig. S11**), they showed acceptable homoscedasticity. Q-Q plots of standardised residuals against the values expected under normal distribution broadly supported the normality assumption, although to some extent they resembled 'light-tailed' distributions (particularly for the leg pain model), where more data is concentrated in the centre of the distribution, and first quantiles occur at larger than expected values and the last quantiles at lower than expected values. Cook's distance values were consistently below 1 (≤0.01 across all outcomes) suggesting no highly influential cases.

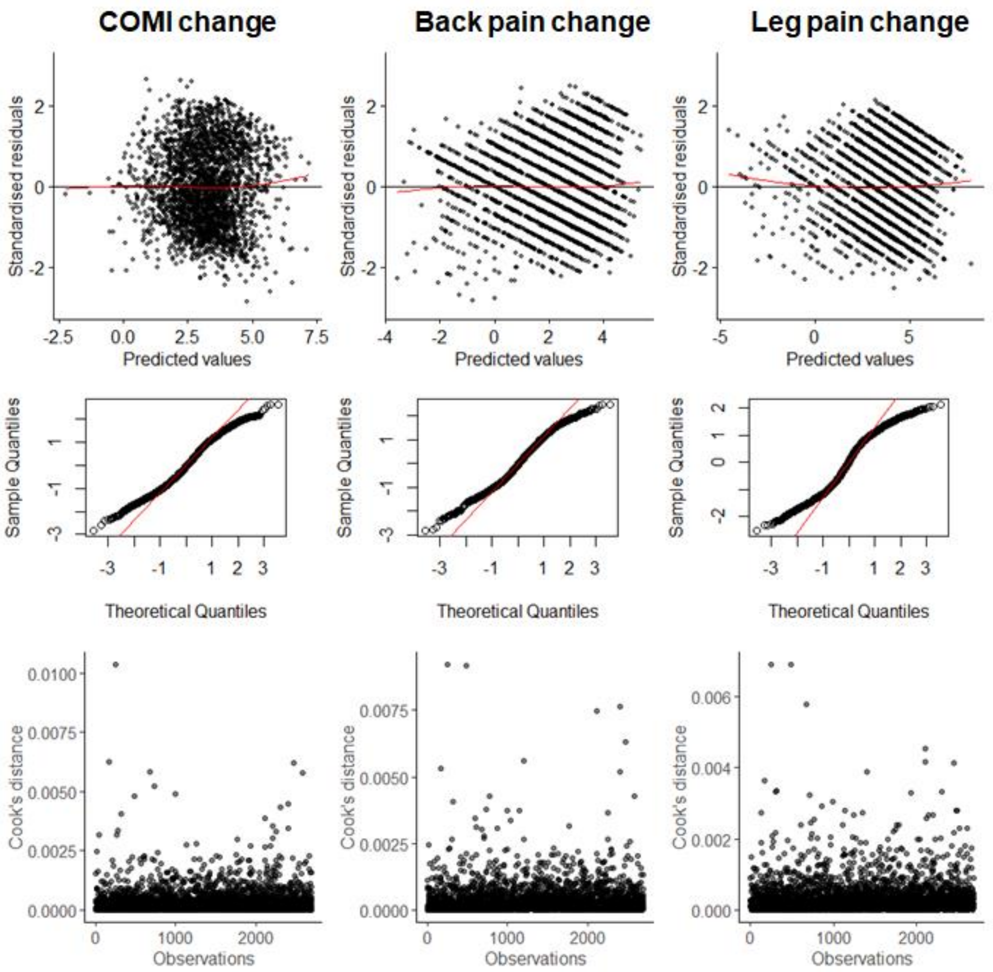

**Fig. S11.** Diagnostic plots for linear regression models on change in Core Outcome Measures Index (COMI), back pain, and leg pain. Standardised residuals are plotted against predicted values of each outcome with best loess fit line in red (top panel), and against values expected under normal distribution with a theoretical normal line in red (middle panel), and Cook's distance is plotted against observation number (bottom panel).

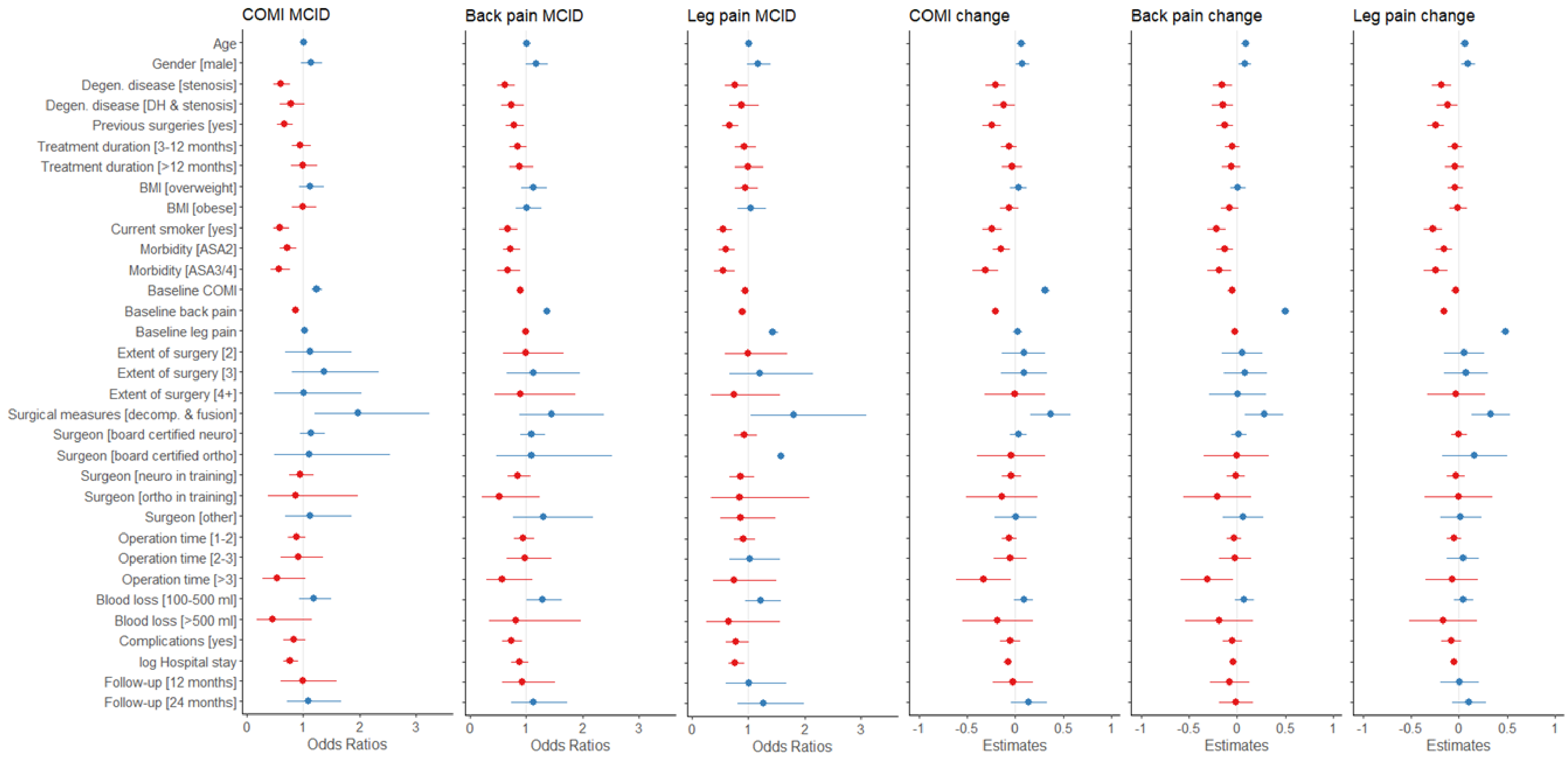

**Fig. S12.** Odds Ratios and standardised regression coefficients (Estimates) with 95% confidence intervals for predictors of Minimal Clinically Important Difference (MCID) and continuous change scores in Core Outcome Measures Index (COMI), back pain, and leg pain intensity. Odds Ratios <1 and Estimates <0 (red) reflect negative predictors, i.e., presence of or higher scores on these factors are associated with lower odds of achieving MCID or less improvement, and Odds Ratios >1 and Estimates >0 (blue) indicate positive predictors, i.e., presence of or higher scores on these factors are associated with greater odds of achieving MCID or more improvement on outcome measures.
ASA, American Society of Anaesthesiologists morbidity class; BMI, body mass index; DH, disc herniation; neuro, neurosurgeon; ortho, orthopaedic surgeon.

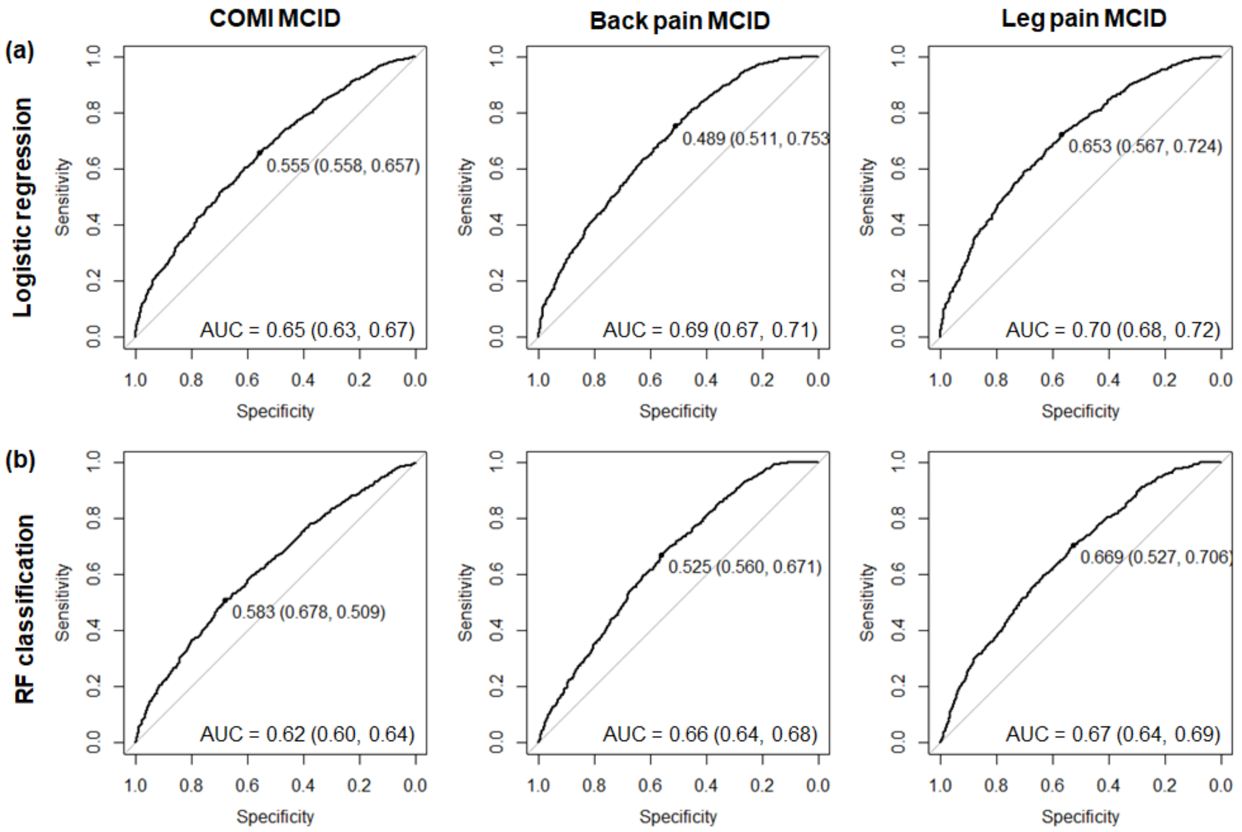

**Fig. S13.** Discrimination ability of the (a) logistic regression and (b) random forest classification models when fitted to the development data for MCID in COMI, back pain, and leg pain. Plots illustrate Receiver-Operating Characteristic (ROC) curves with an optimal probability threshold (black point on the ROC curve; specificity and sensitivity indicated in brackets). Area Under the Curve (AUC) is reported for each ROC with 95% confidence interval.

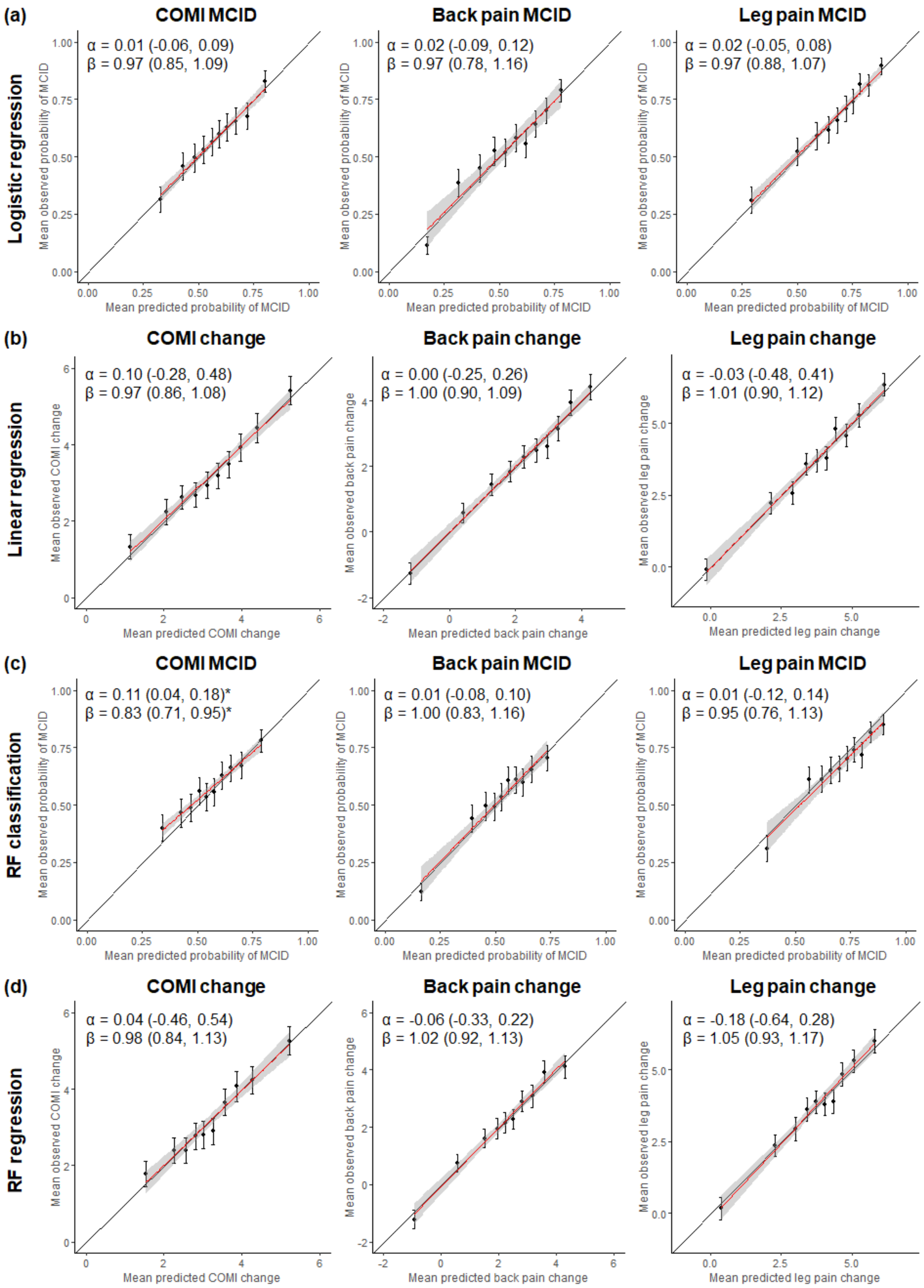

**Fig. S14.** Calibration plots for (a) logistic regression, (b) linear regression, (c) random forest (RF) classification, and (d) RF regression models when fitted to the development data for each outcome. Points represent mean predicted and observed probabilities of MCID (a, c) or change scores (b, d) in each decile with 95% CI of the observed mean. Calibration lines of best fit are plotted in red with 95% CIs in grey, and their intercept (α) and slope (β) estimates with 95% CIs are presented in the top-left corner of each plot. *95% CI of the intercept does not include 0, or the 95% CI of the slope does not include 1, indicating significant deviation from the perfect fit.

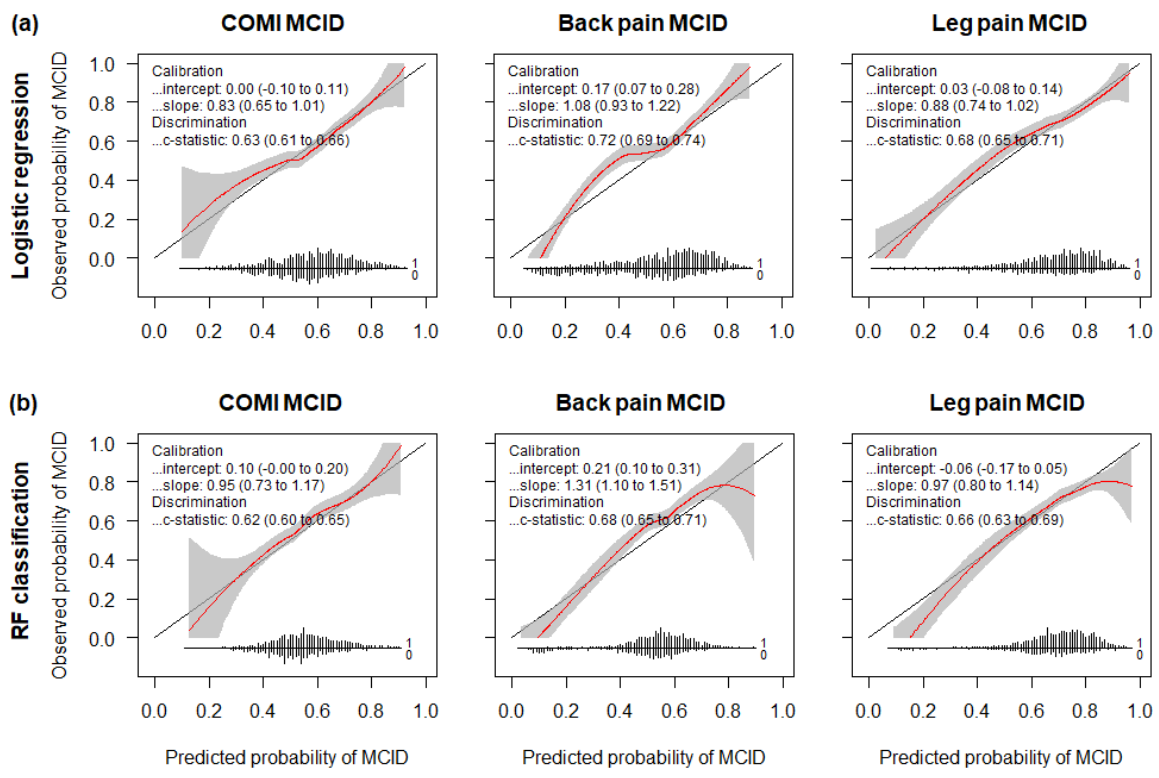

**Fig. S15.** Flexible calibration curves fitted to ungrouped observed vs. predicted MCID probabilities for (a) logistic regression and (b) random forest (RF) classification on the validation data. Loess (span = 0.75) calibration lines are plotted in red with 95% CI in grey, and their intercept and slope estimates, as well as c-statistic corresponding to the area under the curve, are presented in the top-left corner of each plot with 95% CIs. Line of perfect calibration is presented in black as reference.

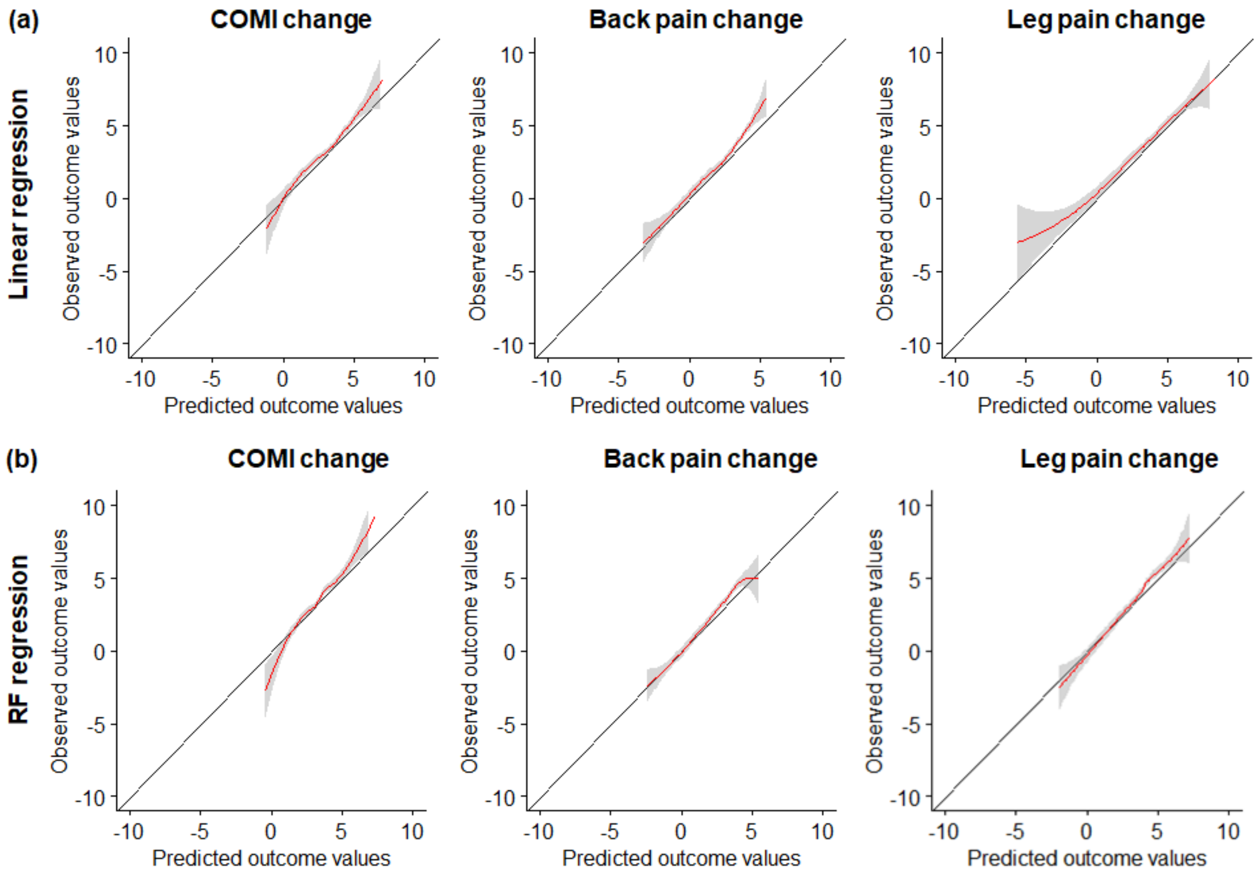

**Fig. S16.** Flexible calibration curves fitted to ungrouped observed vs. predicted change outcome values for (a) linear and (b) random forest (RF) regression on the validation data. Loess (span = 0.75) calibration lines are plotted in red with 95% CI in grey. Line of perfect calibration is presented in black as reference.

#### Results S3: Individual predictions formulae

To make individual predictions based on the developed logistic regression models, for instance, the probability (*p*) of achieving MCID in back pain can be expressed as:

$$p = 1 / (1 + \exp(-(0.01 * age + 0.16 * gender[male] - 0.46 * degen\ disease[stenosis] - 0.3 * degen\ disease[DH\&stenosis] - 0.25 * previous\ surgery[yes] - 0.17 * treatment\ duration[3 - 12m] - 0.12 * treatment\ duration[> 12m] + 0.12 * BMI[overweight] + 0.02 * BMI[obese] - 0.41 * current\ smoker[yes] - 0.32 * morbidity[ASA2] - 0.4 * morbidity[ASA3/4] - 0.1 * COMI + 0.31 * back\ pain - 0.01 * leg\ pain - 0.01 * extent\ surgery[2] + 0.12 * extent\ surgery[3] - 0.1 * extent\ surgery[4] + 0.37 * surgical\ measures[decompression\&fusion] + 0.1 * surgeon[board\ neruo] + 0.09 * surgeon[board\ ortho] - 0.1 * surgeon[neuro\ training] - 0.64 * surgeon[ortho\ training] + 0.27 * surgeon[other] - 0.05 * operation\ time[1 - 2h] - 0.02 * operation\ time[2 - 3h] - 0.55 * operation\ time[> 3h] + 0.25 * blood\ loss[100 - 500ml] - 0.19 * blood\ loss[> 500ml] - 0.31 * complications[yes] - 0.12 * \log(hospital\ stay + 1) - 0.07 * followup[12m] + 0.12 * followup[23m] - 1.29)))$$

where *gender[male]* has a value of 1 if the patient is male, and 0 otherwise; *degen disease[stenosis]* has a value of 1 if the patient has stenosis without disc herniation, and 0 otherwise, *degen disease[DH&stenosis]* has a value of 1 if the patient has disc herniation and stenosis, and 0 otherwise, and both *degen disease* terms have a value of 0 if the patient has disc herniation without stenosis, which is the reference term for this factor in the regression model. 1 or 0 values are assigned to other terms in the equation accordingly (see **Table 1** for reference levels). Probability (*p*) multiplied by 100 would reflect percentage chance that a patient with specified characteristics achieves MCID in back pain. To make individual predictions about the probability of achieving MCID in COMI or leg pain, value of each predictor term should be multiplied by its corresponding log odds as reported in **Table 1**, and model intercept should be included as the final term.

For linear regression models, analogically to the back pain MCID model, individual predictions of change in back pain can be made by multiplying each unstandardised beta coefficient by the value of patient's respective characteristics (1 if category is true, and 0 it is false or consistent with the reference level of a particular predictor) and summing up the score together with the model intercept (see **Table 1**), according to the following equation:

$$predicted = 0.02 * age + 0.28 * gender[male] - 0.51 * degen\ disease[stenosis] - 0.49 * degen\ disease[DH\&stenosis] - 0.43 * previous\ surgery[yes] - 0.17 * treatment\ duration[3 - 12m] - 0.19 * treatment\ duration[> 12m] + 0.03 * BMI[overweight] - 0.26 * BMI[obese] - 0.71 * current\ smoker[yes] - 0.41 * morbidity[ASA2] - 0.6 * morbidity[ASA3/4] - 0.12 * COMI + 0.62 * back\ pain - 0.03 * leg\ pain + 0.18 * extent\ surgery[2] + 0.29 * extent\ surgery[3] + 0.02 * extent\ surgery[4] + 0.93 * surgical\ measures[decompression\&fusion] + 0.07 * surgeon[board\ neuro] - 0.02 * surgeon[board\ ortho] - 0.04 * surgeon[neuro\ training] - 0.68 * surgeon[ortho\ training] + 0.21 * surgeon[other] - 0.11 * operation\ time[1 - 2h] - 0.0 * operation\ time[2 - 3h] - 1.03 * operation\ time[> 3h] + 0.25 * blood\ loss[100 - 500ml] - 0.61 * blood\ loss[> 500ml] - 0.16 * complications[yes] - 0.26 * log(hospital\ stay + 1) - 0.25 * followup[12m] - 0.03 * followup[24] - 0.93)))$$

**Table S5.** Results of the multivariate logistic regression models on MCID outcomes (log-odds) and multivariate linear regression models on continuous outcomes (unstandardised beta coefficients) in the development data.

| | | ***COMI MCID*** | ***Back pain MCID*** | ***Leg pain MCID*** | ***COMI change*** | ***Back pain change*** | ***Leg pain change*** |
|---|---|---|---|---|---|---|---|
| ***Predictors*** | | *Log-Odds (95% CI)* | *Log-Odds (95% CI)* | *Log-Odds (95% CI)* | *Beta (95% CI)* | *Beta (95% CI)* | *Beta (95% CI)* |
| (Intercept) | | -0.68 (-1.57, 0.20) | **-1.29 (-2.20, -0.38)** | -0.72 (-1.67, 0.23) | -0.56 (-1.77, 0.65) | -0.93 (-2.17, 0.30) | -0.59 (-1.97, 0.79) |
| Age (years) | | **0.01 (0.00, 0.02)** | **0.01 (0.01, 0.02)** | **0.01 (0.00, 0.02)** | **0.01 (0.00, 0.02)** | **0.02 (0.01, 0.03)** | **0.02 (0.00, 0.03)** |
| Gender (ref. female) | male | 0.13 (-0.03, 0.29) | 0.16 (-0.00, 0.33) | 0.17 (-0.01, 0.34) | **0.23 (0.01, 0.46)** | **0.28 (0.05, 0.51)** | **0.36 (0.11, 0.61)** |
| Degen. disease (ref. disc herniation) | stenosis | **-0.5 (-0.75, -0.26)** | **-0.46 (-0.71, -0.21)** | **-0.27 (-0.53, -0.01)** | **-0.64 (-0.96, -0.31)** | **-0.51 (-0.84, -0.18)** | **-0.67 (-1.04, -0.29)** |
| | disc herniation & stenosis | -0.25 (-0.51, 0.02) | **-0.3 (-0.57, -0.03)** | -0.12 (-0.41, 0.18) | **-0.37 (-0.73, -0.01)** | **-0.49 (-0.86, -0.13)** | **-0.45 (-0.86, -0.04)** |
| Previous surgeries (ref. no) | yes | **-0.4 (-0.60, -0.20)** | **-0.25 (-0.46, -0.04)** | **-0.41 (-0.62, -0.19)** | **-0.74 (-1.02, -0.46)** | **-0.43 (-0.72, -0.15)** | **-0.89 (-1.21, -0.57)** |
| Treatment duration (ref. <3 months) | 3-12 months | -0.05 (-0.23, 0.14) | -0.17 (-0.35, 0.02) | -0.06 (-0.26, 0.14) | -0.2 (-0.45, 0.05) | -0.17 (-0.43, 0.08) | -0.16 (-0.44, 0.13) |
| | >12 months | 0 (-0.24, 0.23) | -0.12 (-0.35, 0.12) | -0.01 (-0.26, 0.24) | -0.1 (-0.41, 0.22) | -0.19 (-0.51, 0.13) | -0.17 (-0.53, 0.19) |
| BMI (ref. normal) | overweight | 0.12 (-0.08, 0.32) | 0.12 (-0.08, 0.32) | -0.05 (-0.26, 0.17) | 0.11 (-0.16, 0.38) | 0.03 (-0.24, 0.31) | -0.14 (-0.45, 0.17) |
| | obese | 0 (-0.22, 0.21) | 0.02 (-0.20, 0.24) | 0.04 (-0.20, 0.28) | -0.19 (-0.48, 0.11) | -0.26 (-0.56, 0.05) | -0.04 (-0.38, 0.30) |
| Current smoker (ref. no) | yes | **-0.52 (-0.75, -0.28)** | **-0.41 (-0.65, -0.17)** | **-0.57 (-0.82, -0.33)** | **-0.73 (-1.05, -0.41)** | **-0.71 (-1.03, -0.38)** | **-1 (-1.36, -0.63)** |
| Morbidity (ref. ASA1) | ASA2 | **-0.33 (-0.54, -0.13)** | **-0.32 (-0.53, -0.11)** | **-0.5 (-0.73, -0.27)** | **-0.44 (-0.71, -0.16)** | **-0.41 (-0.69, -0.13)** | **-0.57 (-0.89, -0.26)** |
| | ASA3/4 | **-0.55 (-0.85, -0.26)** | **-0.4 (-0.71, -0.10)** | **-0.59 (-0.92, -0.27)** | **-0.94 (-1.35, -0.54)** | **-0.6 (-1.01, -0.19)** | **-0.91 (-1.37, -0.44)** |
| Baseline COMI | | **0.22 (0.15, 0.29)** | **-0.1 (-0.17, -0.03)** | -0.05 (-0.13, 0.02) | **0.63 (0.54, 0.72)** | **-0.12 (-0.21, -0.02)** | -0.08 (-0.18, 0.02) |
| Baseline back pain | | **-0.14 (-0.18, -0.11)** | **0.31 (0.27, 0.35)** | **-0.11 (-0.16, -0.07)** | **-0.24 (-0.28, -0.19)** | **0.62 (0.57, 0.67)** | **-0.21 (-0.27, -0.16)** |
| Baseline leg pain | | 0.02 (-0.02, 0.07) | -0.01 (-0.06, 0.04) | **0.37 (0.31, 0.42)** | 0.04 (-0.03, 0.10) | -0.03 (-0.10, 0.03) | **0.83 (0.76, 0.91)** |
| Extent of surgery (ref. 1) | 2 | 0.13 (-0.38, 0.62) | -0.01 (-0.52, 0.50) | 0 (-0.55, 0.52) | 0.28 (-0.41, 0.96) | 0.18 (-0.51, 0.88) | 0.22 (-0.56, 1.00) |
| | 3 | 0.31 (-0.23, 0.85) | 0.12 (-0.44, 0.67) | 0.19 (-0.40, 0.76) | 0.29 (-0.45, 1.02) | 0.29 (-0.47, 1.04) | 0.28 (-0.56, 1.12) |
| | ≥4 | 0.01 (-0.70, 0.71) | -0.1 (-0.83, 0.63) | -0.29 (-1.04, 0.45) | -0.02 (-0.99, 0.95) | 0.02 (-0.97, 1.01) | -0.11 (-1.22, 1.00) |
| Surgical measures (ref. decompression) | decompression & fusion | **0.68 (0.19, 1.19)** | 0.37 (-0.12, 0.87) | **0.59 (0.06, 1.14)** | **1.14 (0.48, 1.79)** | **0.93 (0.27, 1.60)** | **1.22 (0.48, 1.97)** |
| Surgeon (ref. specialized spine) | board certified neuro | 0.13 (-0.06, 0.33) | 0.1 (-0.10, 0.29) | -0.06 (-0.27, 0.14) | 0.1 (-0.16, 0.36) | 0.07 (-0.20, 0.34) | 0 (-0.30, 0.30) |
| | board certified ortho | 0.11 (-0.70, 0.96) | 0.09 (-0.76, 0.93) | 0.46 (-0.47, 1.53) | -0.12 (-1.22, 0.97) | -0.02 (-1.14, 1.10) | 0.6 (-0.65, 1.85) |
| | neuro in training | -0.05 (-0.28, 0.18) | -0.16 (-0.39, 0.07) | -0.14 (-0.39, 0.10) | -0.12 (-0.43, 0.19) | -0.04 (-0.36, 0.27) | -0.1 (-0.45, 0.25) |
| | ortho in training | -0.14 (-0.97, 0.69) | -0.64 (-1.52, 0.21) | -0.15 (-1.03, 0.77) | -0.42 (-1.57, 0.72) | -0.68 (-1.85, 0.48) | -0.02 (-1.33, 1.28) |
| | other | 0.12 (-0.37, 0.63) | 0.27 (-0.25, 0.79) | -0.14 (-0.66, 0.40) | 0.03 (-0.66, 0.71) | 0.21 (-0.49, 0.90) | 0.08 (-0.70, 0.86) |
| Operation time (ref. <1) | 1-2 | -0.13 (-0.31, 0.05) | -0.05 (-0.24, 0.13) | -0.08 (-0.27, 0.12) | -0.19 (-0.43, 0.06) | -0.11 (-0.36, 0.14) | -0.19 (-0.47, 0.09) |
| | 2-3 | -0.09 (-0.48, 0.30) | -0.02 (-0.42, 0.38) | 0.03 (-0.39, 0.46) | -0.15 (-0.68, 0.39) | -0.06 (-0.61, 0.48) | 0.15 (-0.46, 0.76) |
| | >3 | -0.6 (-1.26, 0.05) | -0.55 (-1.21, 0.11) | -0.29 (-0.98, 0.41) | **-1 (-1.88, -0.13)** | **-1.03 (-1.93, -0.14)** | -0.27 (-1.28, 0.73) |
| Blood loss (ref. <100 ml) | 100-500 ml | 0.17 (-0.06, 0.41) | **0.25 (0.01, 0.49)** | 0.2 (-0.05, 0.46) | 0.27 (-0.05, 0.59) | 0.25 (-0.07, 0.58) | 0.18 (-0.19, 0.54) |
| | >500 ml | -0.76 (-1.72, 0.12) | -0.19 (-1.08, 0.68) | -0.43 (-1.31, 0.46) | -0.55 (-1.70, 0.59) | -0.61 (-1.78, 0.55) | -0.63 (-1.93, 0.68) |

| | | | | | | | |
|---|---|---|---|---|---|---|---|
| Complications (ref. no) | yes | -0.19 (-0.43, 0.05) | **-0.31 (-0.56, -0.06)** | -0.24 (-0.50, 0.01) | -0.15 (-0.48, 0.18) | -0.16 (-0.50, 0.18) | -0.29 (-0.66, 0.09) |
| (log) Hospital stay (days) | | **-0.25 (-0.42, -0.08)** | -0.12 (-0.29, 0.05) | **-0.25 (-0.43, -0.08)** | **-0.41 (-0.64, -0.18)** | **-0.26 (-0.49, -0.02)** | **-0.33 (-0.59, -0.07)** |
| Follow-up (ref. 3 months) | 12 months | -0.01 (-0.49, 0.47) | -0.07 (-0.56, 0.42) | 0.02 (-0.50, 0.52) | -0.06 (-0.71, 0.59) | -0.25 (-0.91, 0.42) | 0.03 (-0.72, 0.77) |
| | 24 months | 0.1 (-0.33, 0.52) | 0.12 (-0.31, 0.55) | 0.25 (-0.21, 0.69) | 0.44 (-0.13, 1.01) | -0.03 (-0.61, 0.55) | 0.39 (-0.26, 1.04) |

*Note.* Coefficients in bold indicate significant predictors (95% CI does not include 0). ASA, American Society of Anaesthesiologists morbidity class; BMI, body mass index; CI, confidence interval; COMI, Core Outcome Measures Index; MCID, Minimal Clinically Important Difference; neuro, neurosurgeon; ortho, orthopaedic surgeon; ref., reference term.

#### Results S4: Sensitivity complete case analysis

Similar to the primary analysis, dataset with participants who had complete data on all predictor variables was divided into the development (N = 1634) and validation data (N = 669) based on the year of surgery (before or from 2017 onwards). The same independent variables were included in the logistic and linear regression models to predict MCID and continuous change in COMI, back pain, and leg pain. As summarised in **Table S5**, the model performance both in the development and validation data was similar or worse in the complete case analysis compared to the primary analysis on the imputed data. Overall, there were no systematic differences in which predictors showed significant prognostic effects, except for the current smoking status, which consistently predicted all outcomes in the primary analysis, but none of the outcomes in the sensitivity analysis.

**Table S6.** Comparison of the results from the primary regression analyses using imputed predictor data and from the sensitivity analyses using only cases with complete predictor data.

| *Predictors* | *COMI MCID* | *Change in COMI* | *Back pain MCID* | *Change in back pain* | *Leg pain MCID* | *Change in leg pain* |
|---|---|---|---|---|---|---|
| | *Significant predictors: imputed data / complete case analysis* | | | | | |
| **Age** | ● / ● | ● / ● | ● / ● | ● / ● | ● / ● | ● / ● |
| **Gender** (male) | | ● / - | | ● / - | | ● / ● |
| **Degen. disease** (stenosis / disc herniation & stenosis) | ● / ● | ● / ● | ● / ● | ● / ● | ● / ● | ● / ● |
| **Previous surgeries** (yes) | ● / ● | ● / ● | ● / - | ● / ● | ● / ● | ● / ● |
| **Treatment duration** | | | | | | |
| **Obesity** | | | | | | |
| **Current smoker** (yes) | ● / - | ● / - | ● / - | ● / - | ● / - | ● / - |
| **Morbidity** (>ASA) | ● / ● | ● / ● | ● / ● | ● / ● | ● / ● | ● / ● |
| **Baseline COMI** | ● / ● | ● / ● | ● / ● | ● / ● | | - / ● |
| **Baseline back pain** | ● / ● | ● / ● | ● / ● | ● / ● | ● / ● | ● / ● |
| **Baseline leg pain** | | | | | ● / ● | ● / ● |
| **Extent of surgery** | | | | | - / ● | |
| **Surgical measures** (decompression & fusion) | ● / ● | ● / ● | | ● / ● | ● / ● | ● / ● |
| **Surgeon** | | | - / ● | | | |
| **Operation time** (>3h) | | ● / - | | ● / - | | |
| **Blood loss** | - / ● | | ● / - | | | |
| **Complications** (yes) | | | ● / - | | | |
| **(log) Hospital stay** | ● / ● | ● / ● | | ● / ● | ● / ● | ● / ● |
| **Follow-up** | - / ● | | | | | |
| *Model performance in development data* (N = 2691 [imputed] / N = 1634 [complete case]) | | | | | | |
| Nagelkerke $R^2$ | 0.10 / 0.10 | | 0.17 / 0.15 | | 0.17 / 0.17 | |
| Adjusted $R^2$ | | 0.13 / 0.12 | | 0.22 / 0.19 | | 0.22 / 0.21 |
| Hosmer-Lemeshow p value | 0.727 / 0. 877 | | 0.003 / 0.041 | | 0.703 / 0.832 | |
| r (observed vs. predicted) | | 0.38 / 0.37 | | 0.48 / 0.46 | | 0.48 / 0.47 |
| AUC (95% CI) | 0.65 (0.63, 0.67) / 0.66 (0.63, 0.68) | | 0.69 (0.67, 0.71) / 0.68 (0.65, 0.70) | | 0.70 (0.68, 0.72) / 0.70 (0.67, 0.73) | |
| *Model performance in validation data* (N = 1616 [imputed] / N = 669 [complete case]) | | | | | | |

| | | | |
|---|---|---|---|
| Hosmer-Lemeshow p value | 0.065 / 0.018 | <0.001 / 0.059 | 0.229 / 0.002 |
| r (observed vs. predicted) | 0.40 / 0.40 | 0.53 / 0.47 | 0.47 / 0.46 |
| AUC (95% CI) | 0.63 (0.61, 0.66) / 0.63 (0.58, 0.67) | 0.72 (0.69, 0.74) / 0.69 (0.65, 0.73) | 0.68 (0.65, 0.71) / 0.66 (0.62, 0.71) |

*Note*. Circles represent significant predictors (blue – positive, red – negative) found in each logistic and linear regression model on the imputed / complete case data. Dashes (‘-‘) indicate that particular predictors which were significant in the primary analysis were not significant in the sensitivity analysis, or vice versa. Model calibration and discrimination metrics from imputed / complete case analyses are presented for the development and validation data.
95% CI, confidence interval; ASA, American Society of Anaesthesiologists morbidity class; AUC, Area Under the Receiver-Operating Characteristic Curve; COMI, Core Outcome Measures Index; MCID, Minimal Clinically Important Difference; r, Pearson’s correlation coefficient.

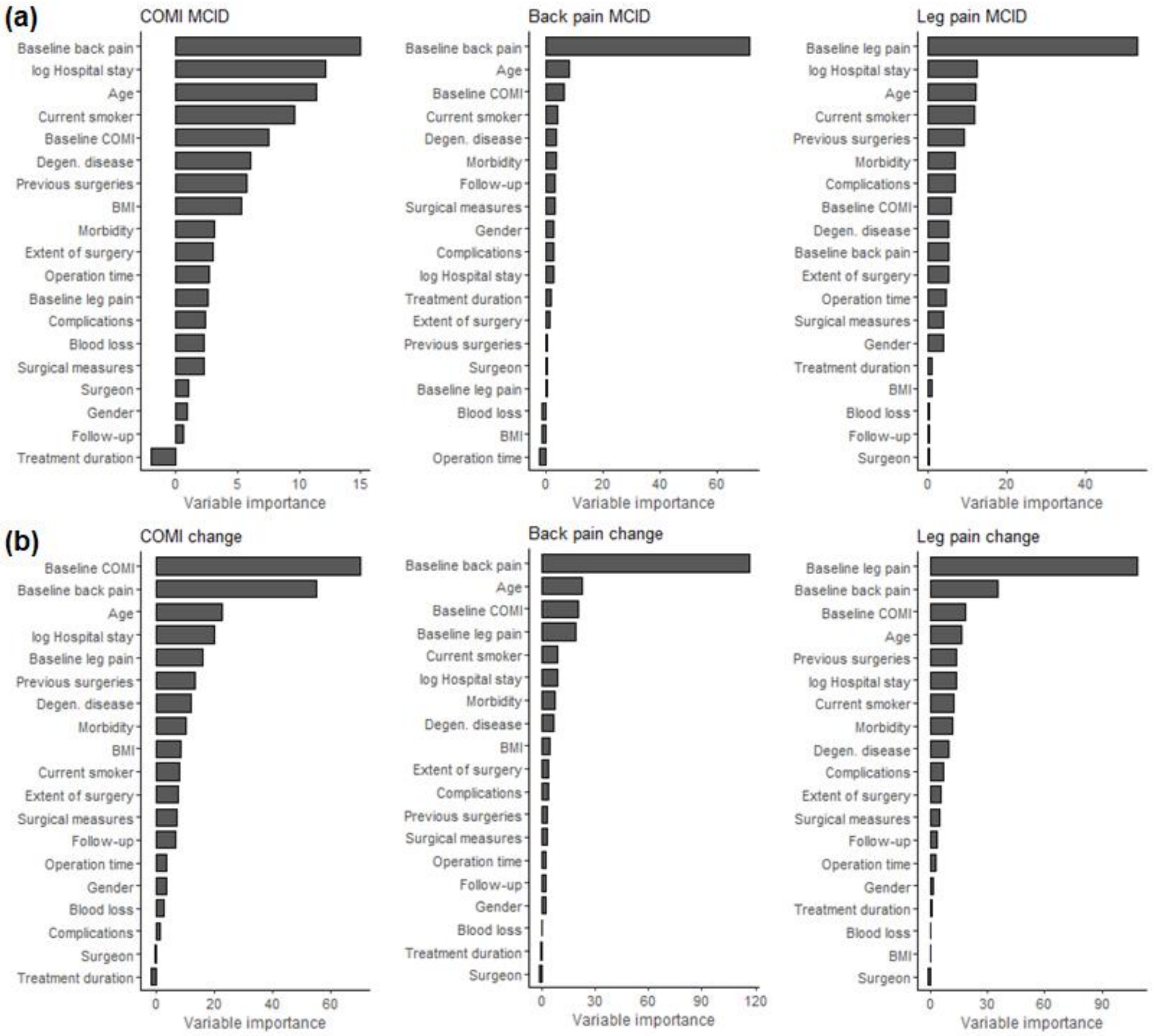

**Fig. S17.** Random forests relative variable importance plots for (a) Core Outcome Measures Index (COMI), back pain, and leg pain Minimal Clinically Important Difference (MCID); and (b) continuous change on these outcomes in the development data. Variable importance was calculated based on mean decrease in classification accuracy (MCID outcomes) and percent increase in mean square error (change outcomes) when a variable was omitted from the training set.

| Section/Topic | Item | | Checklist Item | Page |
|---|---|---|---|---|
| **Title and abstract** | | | | |
| Title | 1 | D;V | Identify the study as developing and/or validating a multivariable prediction model, the target population, and the outcome to be predicted. | 1 |
| Abstract | 2 | D;V | Provide a summary of objectives, study design, setting, participants, sample size, predictors, outcome, statistical analysis, results, and conclusions. | 2 |
| **Introduction** | | | | |
| Background and objectives | 3a | D;V | Explain the medical context (including whether diagnostic or prognostic) and rationale for developing or validating the multivariable prediction model, including references to existing models. | 3-4 |
| | 3b | D;V | Specify the objectives, including whether the study describes the development or validation of the model or both. | 4 |
| **Methods** | | | | |
| Source of data | 4a | D;V | Describe the study design or source of data (e.g., randomized trial, cohort, or registry data), separately for the development and validation data sets, if applicable. | 4 |
| | 4b | D;V | Specify the key study dates, including start of accrual; end of accrual; and, if applicable, end of follow-up. | 4 |
| Participants | 5a | D;V | Specify key elements of the study setting (e.g., primary care, secondary care, general population) including number and location of centres. | 4 |
| | 5b | D;V | Describe eligibility criteria for participants. | 5 |
| | 5c | D;V | Give details of treatments received, if relevant. | 5 |
| Outcome | 6a | D;V | Clearly define the outcome that is predicted by the prediction model, including how and when assessed. | 5-6 |
| | 6b | D;V | Report any actions to blind assessment of the outcome to be predicted. | 5 |
| Predictors | 7a | D;V | Clearly define all predictors used in developing or validating the multivariable prediction model, including how and when they were measured. | 6, Table1 |
| | 7b | D;V | Report any actions to blind assessment of predictors for the outcome and other predictors. | 6 |
| Sample size | 8 | D;V | Explain how the study size was arrived at. | 6, Results S1 |
| Missing data | 9 | D;V | Describe how missing data were handled (e.g., complete-case analysis, single imputation, multiple imputation) with details of any imputation method. | 6-7, Methods S2, Figs.S1-S4, Table S1 |
| Statistical analysis methods | 10a | D | Describe how predictors were handled in the analyses. | 6, Methods S1 |
| | 10b | D | Specify type of model, all model-building procedures (including any predictor selection), and method for internal validation. | 7-10 |
| | 10c | V | For validation, describe how the predictions were calculated. | 8-10 |
| | 10d | D;V | Specify all measures used to assess model performance and, if relevant, to compare multiple models. | 8-10 |
| | 10e | V | Describe any model updating (e.g., recalibration) arising from the validation, if done. | NA |
| Risk groups | 11 | D;V | Provide details on how risk groups were created, if done. | NA |
| Development vs. validation | 12 | V | For validation, identify any differences from the development data in setting, eligibility criteria, outcome, and predictors. | 7, Methods S1 |
| **Results** | | | | |
| Participants | 13a | D;V | Describe the flow of participants through the study, including the number of participants with and without the outcome and, if applicable, a summary of the follow-up time. A diagram may be helpful. | 10-11, Fig. S5 |
| | 13b | D;V | Describe the characteristics of the participants (basic demographics, clinical features, available predictors), including the number of participants with missing data for predictors and outcome. | 10, Table S3, Results S1 |
| | 13c | V | For validation, show a comparison with the development data of the distribution of important variables (demographics, predictors and outcome). | 11, Table S4, Results S1 |
| Model development | 14a | D | Specify the number of participants and outcome events in each analysis. | 10-11, Table1, Table2 |
| | 14b | D | If done, report the unadjusted association between each candidate predictor and outcome. | NA |
| Model specification | 15a | D | Present the full prediction model to allow predictions for individuals (i.e., all regression coefficients, and model intercept or baseline survival at a given time point). | 21, Table1, Table S5, Results S3 |
| | 15b | D | Explain how to the use the prediction model. | Results S3 |
| Model performance | 16 | D;V | Report performance measures (with CIs) for the prediction model. | 11-26; Table1, Table2, Figure1, Figure 2, Fig. S13, Fig. S14, |

| | | | | |
|---|---|---|---|---|
| | | | | Fig. S15, Fig. S16 |
| Model-updating | 17 | V | If done, report the results from any model updating (i.e., model specification, model performance). | NA |
| **Discussion** | | | | |
| Limitations | 18 | D;V | Discuss any limitations of the study (such as nonrepresentative sample, few events per predictor, missing data). | 29 |
| Interpretation | 19a | V | For validation, discuss the results with reference to performance in the development data, and any other validation data. | 26-28 |
| | 19b | D;V | Give an overall interpretation of the results, considering objectives, limitations, results from similar studies, and other relevant evidence. | 26-30 |
| Implications | 20 | D;V | Discuss the potential clinical use of the model and implications for future research. | 29-30 |
| **Other information** | | | | |
| Supplementary information | 21 | D;V | Provide information about the availability of supplementary resources, such as study protocol, Web calculator, and data sets. | 31 |
| Funding | 22 | D;V | Give the source of funding and the role of the funders for the present study. | 32 |

*Items relevant only to the development of a prediction model are denoted by D, items relating solely to a validation of a prediction model are denoted by V, and items relating to both are denoted D;V. We recommend using the TRIPOD Checklist in conjunction with the TRIPOD Explanation and Elaboration document.
